# Investigating brain haemodynamics during hypoglycaemia in very preterm neonates using diffuse optical tomography

**DOI:** 10.1101/2025.09.01.25334829

**Authors:** Guy A. Perkins, Silvia Guiducci, Giulia Res, Federica Savio, Daniele Trevisanuto, Elena Priante, Eugenio Baraldi, Alfonso Galderisi, Sabrina Brigadoi

## Abstract

Very preterm neonates are more prone to experience hyper and hypo-glycaemia after birth. To date, there is no available evidence on the local brain hemodynamic response to these glycemic changes. This study uses continuous glucose monitoring (CGM) and diffuse optical tomography (DOT) to investigate this issue. Sixty very preterm neonates were recruited and continuously monitored after birth with CGM and DOT for several days. Patients with only mild (Sensor glucose concentration (SGC) ≥ 48 mg/dL & ≤ 72 mg/dL) or severe (SGC ≤ 47 mg/dL) hypoglycaemia (N=17) were then selected for analysis. DOT data were reconstructed in three 10-minute windows throughout each hypoglycemic event: at the beginning of the event, after the minimum of hypoglycemia, and at the end of the event. Correlations and spatial consistencies were found between changes in blood volume and SGC during these time windows, suggesting a coupling between SGC and brain hemodynamics in very preterm newborns after birth.

## 1 Introduction

It has been estimated that in the US alone during 2015, around 1.6% of all live births were very preterm (≤ 32 weeks gestational age (GA)) and around the world 1 in 10 babies are born prematurely (Barfield, 2018).Very preterm neonates are more prone, compared to full-term newborns, to experiencing hyper (Angelis et al., 2023) and hypo-glycemia after birth due to the impairment of glucose homeostasis characterizing this early age of life. A study of low birth weight preterm neonates found that 80% of patients exhibited hyperglycemia and for small for gestational age (SGA) neonates (Beardsall et al., 2010), the incidence of hypoglycemia has been estimated around 73% (Duvanel et al., 1999). Impaired glucose control in this population has been associated with higher mortality and morbidity as well as with poor neurological outcome (Lucas et al., 1988 and Beardsall, 2005).

The magnitude and breadth of the impact of hypoglycemia on neurodevelopment is still, however, controversial. A review (Shah et al., 2019) found that neonatal hypoglycemia was not associated with neurodevelopmental impairment in early childhood, but it was associated with visual-motor impairment and executive dysfunction. In mid childhood, neonatal hypoglycemia was instead associated with neurodevelopmental impairment and lower literacy and numeracy scores. Another meta-analysis reported that there seems to be an association, although of low quality, between neurodevelopmental outcome and hypoglycemia at birth (Diggikar et al., 2024). Furthermore, a study involving 528 neonates with hypoglycemia (McKinlay et al., 2015) found that when treatment was provided to maintain a sensor glucose concentration of at least 47 mg/dL, the severe hypoglycemia threshold, then hypoglycemia was not associated with an adverse neurologic outcome, which highlights how crucial monitoring severe hypoglycemia is.

Hyperglycemia has been found to be associated with neurological and neurodevelopmental impairments as well, although evidence is still controversial. It has been reported that very preterm newborns experiencing hyperglycemia at birth had a lower white matter volume at 2.5 years (Naseh et al., 2022). Furthermore, an association between hyperglycemia and lower Bayley-III cognitive and motor scores was found in those children born ≤ 28 weeks GA. These results were confirmed by a review (Guiducci et al., 2022), which found an association between hyperglycemia and neurological delay, in particular motor functions, in the first two years of life, which was maintained into childhood. However, the quality of this evidence was reported to be poor.

Most of the reviews on the association between hypo- or hyperglycemia and neurodevelopmental outcome highlight the need for more studies to better characterize this association in a more systematic way. Most of the studies investigating the association of neurodevelopmental outcome and glucose variability relied on a sparse sampling of the glucose concentration in the first days of life, e.g., every 6 hours (Lucas et al., 1988). Another review (Shah et al., 2019) reported that several studies measure glucose concentration on a frequency of once every 24 hours to once every 4 hours. Clinical practice in most hospitals in the world for the preterm population requires glucose concentration to be measured approximately twice a day with heel-prick sampling (Şimsek et al., 2018). This might suggest that the controversial results found in these reviews might be due to an underestimation of the glucose variability and number and duration of hypo or hyperglycemic events at birth due to the lack of knowledge of the glucose concentration during the whole day.

One solution to this problem could be to use continuous glucose monitoring (CGM) at birth (Beardsall et al., 2021), which involves the use of a minimally invasive device able to continuously measure glucose concentration in the interstitial fluid. CGM is widely used in the diabetic population and several clinical trials have demonstrated its safety also in the preterm population (Galderisi et al., 2017, Tottman et al., 2018 and Kalogeropoulou et al., 2023). It has been demonstrated that the use of CGM can reduce glucose variability and time spent outside of euglycemia in preteterms (Galderisi et al., 2017) and therefore achieve a potentially tighter glucose control (Tottman et al., 2018). However, up to date, CGM has not yet obtained regulatory approval to be used in the neonatal population, therefore its use is limited to clinical trials.

Another reason why controversial results were found in reviews linking glucose variability with neurodevelopmental outcome might be the high variability in neurodevelopmental outcome usually reported within the same patients’ group without covariates able to explain this variance (Rasmussen et al., 2020). There is little evidence for what the optimal individual target glucose levels at birth should be (Hay et al., 2009); therefore, glucose adjustments are based on standardized operational thresholds. The variability reported in neurodevelopmental outcome might be explained, therefore, by different individual susceptibility to glucose changes that are not solved by the operational thresholds, leading to different brain areas impacted by this change in glucose concentration in different newborns. This demonstrates the need to investigate how the preterm brain responds during hypo or hyperglycemic events and whether this brain response is linked to later neurodevelopmental outcome, as few studies investigated the association between hypo or hyperglycemic changes and brain responses.

Positron emission tomography (PET) was used on a cohort of preterm neonates to measure cerebral blood flow (CBF) during hypoglycemia (Pryds et al., 1988). They demonstrated a significantly increase of CBF during the beginning of the hypoglycemia, which they suggest could be a compensatory response to support cerebral metabolism during hypoglycemia. The same authors (Pryds et al., 1990), in a following study, further confirmed this inverse relationship between CBF and sensor glucose concentration (SGC) at the start of hypoglycemia, such that neonates with lower SGC had larger CBF, and viceversa. They also found that after 30 minutes of intravenous glucose treatment in hypoglycemic neonates, CBF decreased by an average of 11.3% but was still 37.5% higher than CBF in control neonates. PET, however, requires radioactive tracers and therefore is not a suitable technique nowadays for preterms if not clinically required. Cot-side non-invasive brain imaging methods should be preferred.

Diffuse optics uses near infrared light to non invasively sample the cortical surface with sources and detectors of light placed on the surface of the scalp. Each source-detector couple at a reasonable source-detector distance is a measurement channel. The applied method of this is called near infrared spectroscopy (NIRS) (Jöbsis, 1977), which uses changes in the intensity of the measured light to recover changes in oxygenated (HbO) and deoxygenated (Hb) hemoglobin. NIRS can be a single-channel (Xu et al., 2024) or a multi-channel approach (De Felice et al., 2024), and the outcome is an estimate of the brain hemodynamic response pattern in each channel. Diffuse optical tomography (DOT) (Eggebrecht et al., 2014), instead, uses high-density NIRS to reconstruct images of changes in HbO and Hb on the cortical surface, thus yielding a more spatially detailed picture of the brain response to a specific event.

Few studies investigated the global brain hemodynamic response in newborns to glycemic variability using NIRS. One study measured changed in total hemoglobin concentration (the sum of HbO and Hb, a surrogate of cerebral blood volume (CBV)) before and during an intravenous bolus of glucose infused into preterm neonates experiencing hypoglycemia, using a four-channel NIRS device (Skov and Pryds, 1992). They found that CBV started to decrease at the end of the infusion and that newborns with an initial lower SGC had the highest decrease in CBV, which was in agreement with prior studies using PET (Pryds et al., 1988 and Pryds et al., 1990). In another study, preterm and full-term newborns were monitored with single-channel NIRS immediately after birth (Matterberger et al., 2018). Significant correlation between both cerebral regional oxygen saturation (crSO_2_) and cerebral fractional tissue oxygen extraction (cFTOE) and SGC were found, stronger for the preterm than the full terms populations. In particular, higher SGCs were associated with smaller crSO_2_, and higher cFTOE. No study so far, to our knowledge, has investigated the hemodynamic brain response to hypoglycemia opting for a multi-channel approach, able to yield information on regional hemodynamic changes in different cortical areas.

The BabyGlucoLight clinical trial (Brigadoi et al., 2022) was designed to investigate brain hemodynamic changes associated with hypo and hyperglycemia by continuously monitoring preterms in the first days after birth with DOT and CGM. We have previously demonstrated that continuous long-term monitoring with DOT is safe in preterm newborns, reporting DOT data recorded continuously for 7 days on a 28 weeks GA newborn in the neonatal intensive care unit (Galderisi et al., 2016). Furthermore, newborns of the BabyGlucoLight clinical trial will be followed-up at 12 and 24 months of corrected age to evaluate their neurodevelopment and investigate its association with brain hemodynamic responses to glycemic events at birth.

The aim of this paper is to address the first question that led to the implementation of the BabyGlucoLight clinical trial, i.e., to confirm the association between hypoglycemia and brain hemodynamic changes, as reported in previous studies, and to investigate with DOT whether the hemodynamic response to hypoglycemia varies spatially across newborns and whether it is spatially consistent or not within the same newborn.

## 2 Methods

### 2.1 Patient recruitment

This study is part of a collection of experiments conducted for the BabyGlucolight clinical trial (NCT04347590).

Inclusion crtiera were that neonates had a GA of ≤ 32 weeks or a birth weight of ≤ 1500 g when admitted to the NICU and that they were available for monitoring within the first 48 hours of life from birth. Neonates were excluded if their birth weight was ≤ 500 g, or if there were congenital malformation, perinatal maternal infections, and albinism. Written informed parental consent was obtained before recruitment.

Patients were monitored with either blinded or unblinded CGM while simultaneously monitoring their brain hemodynamics using DOT. Blinded CGM means that whilst patient SGC is measured continuously, clinicians will not have access to measured data, whereas for unblinded patients, clinicians will have access to all SGC measurements. Patients are followed-up at 1 and 2 year of corrected age to evaluate their neurodevelopment.

The trial was approved by the Institutional Ethics Committee of the University Hospital of Padua (Italy) (4773/AO/19 AOP1813 and 5005/AO/21 AOP2216) and conducted according with the Declaration of Helsinki. Participants were enrolled from March 2020 to June 2023 at the Neonatal Intensive Care Unit (NICU) of the University Hospital of Padua. The enrollment was delayed due to the pandemic restrictions in place in our Country.

In total 60 preterms were recruited. For 8 patients the SGC data were missing due to technical problems with the CGM, leaving 52 patients available for analysis (mean gestational age: 30 ± 3 weeks; mean birth weight: 1281 ± 369 grams). In this manuscript, we will be looking at patients who exhibited only hypoglycemic events, as described in the following sub section. Table 1 reports information on the patients enrolled.

**Table 1:** Clinical information of the analyzable patients (N=52) and the S-m-hypo only subgroup (N=17). Values are expressed as median (iqr 1 - iqr 3), or n (%). Sex is expressed as n male:n female.

|  | All patients (N=52) | S-m-hypo subgroup (N=17) |
| --- | --- | --- |
| Neonates |  |  |
| Gestational age (wk) | 30 (29-31) | 30 (29-31) |
| Birth wt (g) | 1300 (1040-1485) | 1380 (1200-1530) |
| Small for gestational age, n (%) | 7 (13%) | 1 (6%) |
| Twins, n (%) | 16 (31%) | 5 (29%) |
| Sex (male:female) | 26:26 | 5:12 |
| CRIB score | 1 (0-3) | 1 (0-1) |
| Mothers |  |  |
| Maternal diabetes, n (%) | 13 (25%) | 5 (29%) |
| PPROM, n (%) | 14 (27%) | 4 (24%) |

### 2.2 Sensor Glucose Concentration acquisition

A Medtronic Guardian 3 device was used (Medtronic, USA) for CGM, giving SGC measurements every 5 minutes (sampling frequency of 0.083 Hz). This device has been validated as safe and reliable to use in this population (Bonet et al., 2025). It should be noted that these measurements are sensor glucose concentrations (SGC), since the sensor measures interstitial fluid GC and estimates blood glucose concentration based on models developed by the manufacturer. The device was placed on the lateral side of the thigh after disinfection (figure 1 A.). The CGM device was placed on the neonate within the first 48 hours of life and was aimed to sample data continuously for 7 days. The device was removed and stopped earlier if clinical care required its removal. The device was calibrated as per the manufacturers guidelines. SGC data was saved to an Excel spreadsheet, which was retrospectively read and processed on Matlab (2023a, Mathworks, USA). SGC data were analyzed and segmented into 5 different event types: severe hyperglycemia (S-hyper, ≥ 180 mg/dL), mild hyperglycemia (m-hyper, 179 to 144 mg/dL), mild hypoglycemia (m-hypo, 72 to 48 mg/dL), severe hypoglycemia (S-hypo, ≤ 47 mg/dL) and euglycemia (143 to 73 mg/dL). Hypo- and hyperglycemic events were analyzed and classified based on the following procedure. The start of a glycemic event was defined as the first four consecutive samples (15 minutes) in euglycemia with a SGC range ≤ 15 mg/dL going backward from the SGC minimum/maximum value. Analogously, the end of a glycemic event was defined as the first four consecutive samples in euglycemia with a SGC range ≤ 15 mg/dL after the minimum/maximum SGC value for that event. For an event to be classified as severe (hyper or hypo), there must be a minimum of three consecutive SGC measurements within the severe threshold. Finally, in some readings, there were gaps between SGC samples. This was primarily due to the fact that the device needed calibrating approximately every 12 hours and if the calibration was not performed, the SGC data would not be recorded until the device was calibrated again. In this case, any gaps in the SGC data were interpolated by taking the average value of SGC from the start and end of the gap. If an event had a gap of more than 10 minutes (3 samples) within it, the event was not considered for analysis due to the unknown SGC within the gap duration. It should also be noted that the minimum reading of the SGC device was 40 mg/dL.

**Figure 1:**
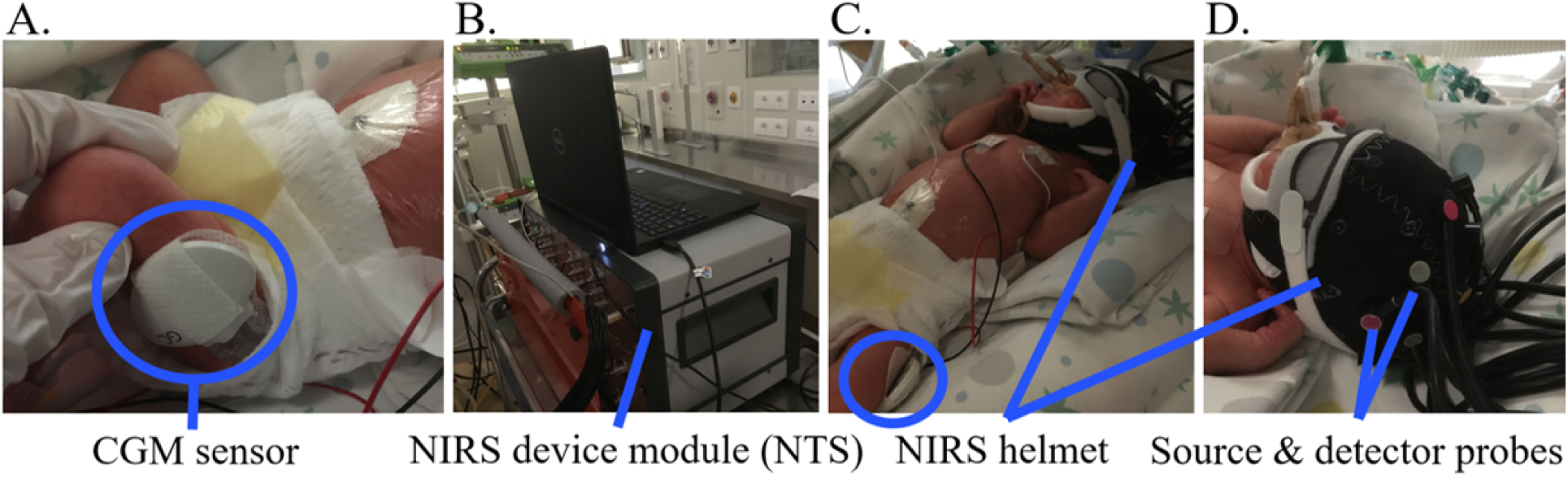
Experimental set up on the preterm. A. The CGM sensor on the preterm thigh. B. The NIRS device module of the NTS (Gowerlabs). C. and D. The NIRS helmet and source-detector probes on the preterm head.

### 2.3 NIRS data acquisition

Table 2 shows the parameters of the device used for NIRS data acquisition. The NIRS data was collected by the clinicians in the hospital, whereby the NIRS cap was placed on each preterm and an automatic source calibration took place before data acquisition (figure 1 B. C. and D.). The NIRS device was tested to be safe on preterms for long continuous acquisitions (order of days) in a prior study (Galderisi et al., 2016). The target duration of continuous NIRS acquisition was the same as the CGM device; however, the acquisition could be stopped and restarted later on if the preterm had to be manipulated or moved for clinical care. The NIRS data were manually synchronized with the SGC data and only the NIRS data synchronized with hypoglycemic events were then further analyzed. Sources and detectors were placed to cover most of the cortical layer, in specific 10-5 positions (figure 3).

**Table 2:** The parameters of the device used for NIRS data acquisition.

| Parameter | Value |
| --- | --- |
| Device Name | NTS Gowerlabs (London, UK) |
| Number of sources | 8 |
| Number of detectors | 8 |
| Max Number of channels | 64 (each $\lambda$) |
| Wavelengths (nm) | 780, 850 |
| Sampling Frequency (Hz) | 10 |
| Laser specification | Class I |

### 2.4 NIRS data preprocessing

For each hypoglycemic event, NIRS data was partitioned into three 10 minute time windows for analysis, namely the first 10 minutes of the event (time window A), from 2 minutes before to 8 minutes after the minimum SGC of the event (time window D) and from 2 minutes before to 8 minutes after the starting point of the final baseline of the event (time window F) (figure 2). This was done because it was not feasible to analyze the entire time series of a hypoglycemic event, which could last hours, using an initial common baseline period, due to the amount of motion artifacts across the event, which could create several baseline shifts.

**Figure 2:**
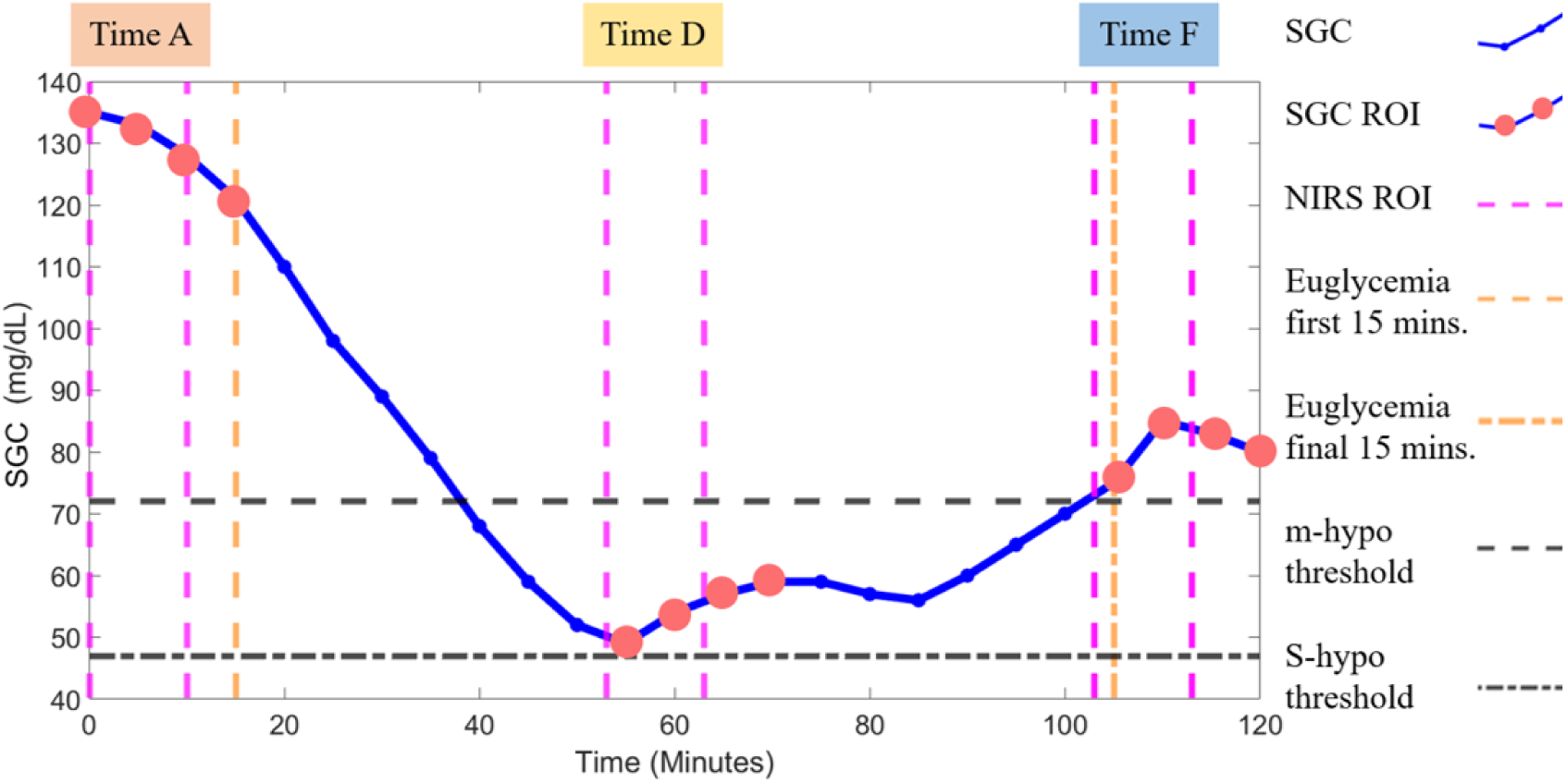
An exemplary mild hypo glycemic event with three regions of interest marked by pink dashed lines to denote three time windows of NIRS analysis, A, D and F. The pink dots denote the SGC samples used for these time windows in the correlation analyses.

**Figure 3:**
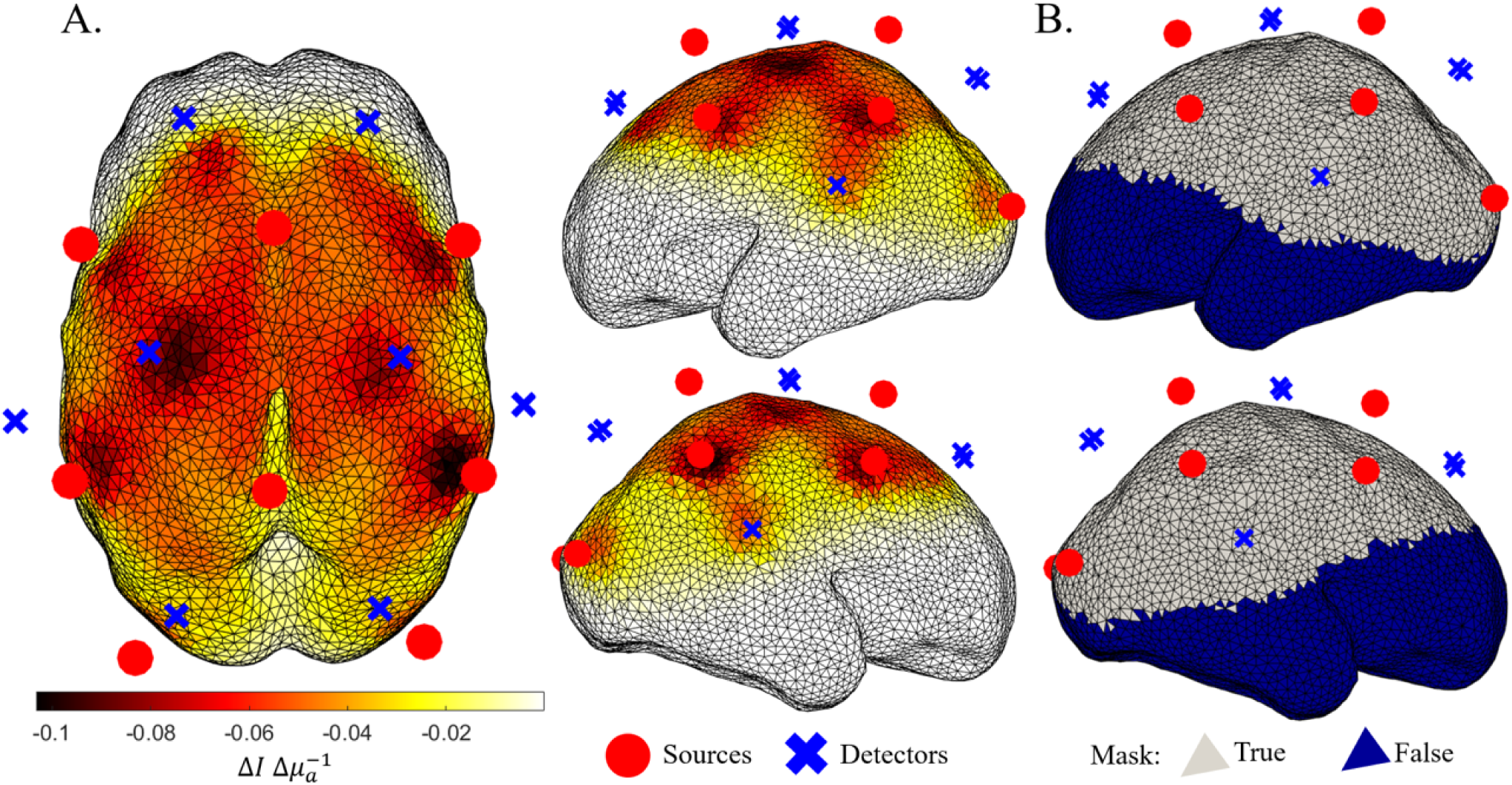
A. The source-detector array over the 30-week GM smooth surface mesh (Brigadoi et al., 2014) with an example Jacobian displayed. B. The coverage threshold mask, showing the nodes (Mask = True) considered for further analysis.

For each 10 minute segment, for each event, signal quality controls were performed on the raw NIRS data to identify good channels. This was done by flagging good channels as those with a mean intensity between 5E-4 to 3 units (manufacturers recommended thresholds) and having SNR ≥ 2 across each 2 minute segment of the 10 minute time window. Furthermore, channels with a change ≥ 0.15 units of intensity were removed, as well as channels with peaks in their frequency spectrum at specific frequencies (0.0667 Hz, 0.0516 Hz and 0.0150 Hz), due to detector saturation. Significant peak values at these frequencies were estimated as a peak power value greater than 1.5 the standard deviation of the mean power values of all good channels. Any event with ≤ 23 good channels across both wavelengths was discarded.

After computing changes in optical density, motion correction was applied. Firstly, due to preterms moving whilst asleep or being manipulated for clinical care, there may be temporary changes in the coupling of the source-detector probes on the scalp, which can give a series of motion artifacts (MA) within seconds of each other or a few seconds of artefactual signal. These instances will be called MA ’trains’ and they were identified using the global variance of temporal derivatives (GVTD) approach (Sherafati et al., 2020). GVTD calculates the global change in Δ*OD* between each time point across all good channels to form a distribution of changes, and for time points where these changes are more than N *σ* above the mean, these time points are labeled as MA. GVTD was applied twice. The first run was used only to identify channels displaying several MAs or MA trains; the second run was performed excluding these highly contaminated channels, with the aim to more precisely detect the artifactual time points.

During the first run, *σ* was set to 4 to label MAs, and an MA train was defined when at least 20 samples were labeled as MA within a 200 sample sliding window sliding along the 10 minute recording. MA trains were then joined together if they occurred within 100 frames of each other. If a channel had a number of MAs greater than the mean ± one standard deviation of MAs across all channels it was unlabeled as a good channel for the GVTD process only. The second pass of GVTD was then performed using *σ* = 4 as before, on an updated set of good channels to identify the MAs and MA trains.

Once the MA trains were found, they were corrected by using a linear interpolation between the start and end of the MA train using the ’pchip’ function (interpn); noise as added to the interpolated signal (0.8 times the mean standard deviation of the signal in the 20 frames before the MA train times by a pseudo-random number between −1 and 1). The aim of the interpolation was to correct MA trains without removing the data points, such that more events could be used for analysis. In the events considered for analysis, MA trains were classified on average in 20.2% of frames per event, with an average of 2.53 MA trains detected per event. Any event with more than 50% of frames classified as MA trains was discarded.

After MA trains were corrected, MAs not defined as trains were corrected by applying the following MA detection and correction techniques. Firstly, a modified version of the Homer2 (Huppert et al., 2009) function hmrMotionArtifactByChannel was used to identify MAs (tMotion = 1.0, tMask = 2, SDThresh = 7 and AmpThresh = 0.1). This function detects MAs by running a sliding window of width tMotion across the signal, to find changes in amplitude greater than AmpThresh or in standard deviation greater than SDThresh. It should be noted that a modification was used to calculate SDThresh (Yang et al., 2022). Then a Spline interpolation with a Savitzky Golay filter was used (hmrMotionCorrectSplineSG) with a frame size of 10, followed by a wavelet filter (hmrMotionCorrectWavelet) with an iqr = 1.1 (Di Lorenzo et al., 2019). A lowpass filter with cut-off frequency of 0.0067 Hz was then applied.

Baseline correction was then applied to the MA corrected Δ*OD*, with the mean of the first 2 minutes of the 10 minute segment used for baseline subtraction. Outlier channels (channels that deviated ≥ 2 *σ* from the mean ΔOD of all good channels for more than 25% of the time points) were removed. Only to remove further outliers, spectroscopic changes in chromophore concentration were calculated using the modified Beer-Lambert law, with an age-specific differential pathlength factors (Scholkmann and Wolf, 2013) and outliers removed, using the same criteria as removing the Δ*OD* outliers. This two-step outlier removal was performed in order to remove channels that may have passed through the MA detection and correction process but then exhibited non realistic changes in Δ*OD* and/or Δ*HbO*. The average percentage of good channels (100% = 128 channels) classified after initial signal pruning was 47.5%, and then 33.9% after outlier removal. Overall, the average number of good channels after all preprocessing was 40, with a minimum of 12 and a maximum of 68. Finally, the Δ*OD* signal was downsampled from 10 Hz to 1 Hz and used to perform image reconstruction.

### 2.5 Image reconstruction

Image reconstruction was performed using NIRFAST (Dehghani et al., 2008) and custom-made code. The 30-week head model from the 4D neonatal head model atlas (Brigadoi et al., 2014) was used as head model for all patients, since the average GA of our patients was 30 weeks. The jacobian was computed with NIRFAST, using default parameters (Figure 3A.). Optical properties were assigned to each tissue type (Uchitel et al., 2023), and are shown in table 3.

**Table 3:** Optical properties used for the head model and jacobian generation. These were taken from Uchitel et al., 2023, for 850nm.

| Layer | $\mu_a$ (mm<sup>-1</sup>) | $\mu'_s$ (mm<sup>-1</sup>) |
| --- | --- | --- |
| Scalp | 0.020 | 0.751 |
| CSF | 0.004 | 0.300 |
| GM | 0.019 | 0.673 |
| WM | 0.021 | 1.011 |
| Brainstem | 0.021 | 1.011 |
| cerebellum | 0.021 | 1.011 |

The inverse problem was solved as previously described in Perkins et al., 2022, using the Moore-Penrose pseudoinverse and Tikhonov regularization of the Jacobian, with a regularisation parameter of 0.5 and beta values of 0.01 used for voxel and data magnitude normalization. To resolve for changes in chromophore, extinction coefficients were used from the GetExtinctions function (Huppert et al., 2009). Volumetric changes in chromophore concentration were then mapped to the GM surface mesh, by assigning the average value of all the volumetric mesh nodes within a 3 mm radius to each GM surface node. A binary mask was created to isolate GM surface nodes sensitive to the array employed in this study. Only nodes within the mask were submitted to further analyses. The mask was created by considering the full-channel Jacobian mapped to the GM surface mesh and selecting nodes with a sensitivity ≥ a coverage threshold (Brigadoi et al., 2018):

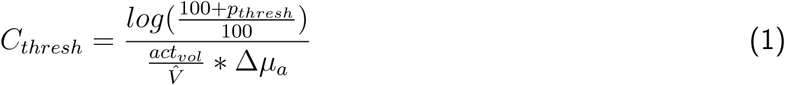

with *p_thresh_* the minimum change in the intensity of the signal that can be measured (1%), *act_vol_* an approximate volume over which a hemodynamic response can be expected to occur (10^3^), *V*^^^ the median Voronoi volume across nodes of the GM mesh (≈ 1cm^3^) and Δ*µ_a_* the approximate change in absorption coefficient expected during a hemodynamic response (approximately 10% of *µ_a_* ≈ 0.001 mm^−1^). The resulting mask is displayed in figure 3B.

To ensure an event had sufficient coverage across the cortex, 9 region of interests (ROIs) were segmented, each centered around 9 EEG 10-20 landmark positions (F3, FZ, F4, C3, CZ, C4, P3, PZ and P4) with a radius of 18 mm. For a given event, if less than 33% of nodes in any ROI had a sensitivity below the coverage threshold, that event was discarded.

### 2.6 Statistical analysis

Changes in HbT, as a proxy for changes in cerebral blood volume, were correlated with the SGC data. In particular, for a given hypoglycemic event, several parameters were calculated from Δ*HbT* in the 10 minute window and from the SGC values in the corresponding 15 minute window (figure 2). The NIRS time window was chosen to be 10 minutes to minimize the chance of large MAs, several MA trains or poor signal quality, whereas the SGC time window was chosen to be 15 minutes such that four samples of SGC could be considered as to provide more meaningful interpretation of SGC statistics. The metrics used for the correlation analysis are reported in table 4. For the SGC metrics, metrics were computed both at the local scale (within the 15 minute window) and at the global scale (considering the entire hypoglycemic event). It should be noted that, since HbT can oscillate within the NIRS time window, we chose to take the absolute value for the HbT range, so that the HbT range metric depicts the actual range, whatever the direction of the changes. Instead, the SGC range was chosen to be kept either positive or negative, since SGC typically either increases or decreases for a specific type of event (hypoglycemia) or within a small time window. The HbT maxima is the maximum value of the absolute change in HbT, and the T value was obtained by computing the Student’s t-test. For HbT, the t-test was computed by comparing how HbT changed compared to a zero meaned change across the 10 minute time window and the t-test for the SGC was computed by comparing how SGC changes with respect to the first SGC value, which we considered as baseline.

**Table 4:** The different metrics calculated for the SGC and HbT data. L denotes ’local’ changes, i.e within the SGC or NIRS ROI (. **figure 2) and G denotes ’global’ changes, i.e across the entire time of the glycemic event).**

| Data Type | Metrics |  |  |  |  |
| --- | --- | --- | --- | --- | --- |
| SGC | L G Variance | L G Mean | L G Range | G Minima | G T-value |
| HbT | L Variance | L Mean | L Range | L Maxima | L T-value |

Linear correlations (R^2^ values) were calculated between the HbT and SGC metrics using the robustfit and corr functions in Matlab, giving 40 direct comparisons of metrics (5 HbT metrics x 8 SGC metrics) for each time window for a given group of patients, for each node on the surface of the cortex. Multiple comparison correction was performed using the false discovery rate (FDR) correction (Benjamini and Hochberg, 1995 and Groppe, 2025).

In addition to these metrics, to evaluate the spatial consistency of the HbT changes within and across patients, the net T-value was calculated either considering all events of the same patient (for newborns having more than 3 events) or all events of all patients. The net T-value was obtained for each node, by subtracting the number of events with a statistically significant negative T-value for HbT (compared to a zero change) from the number of events with a statistically significant positive T-value. Then the net T-value was then normalized by dividing the so obtained value by the total number of glycemic events for the single patient or group of patients being considered.

## 3 Results

### 3.1 SGC Classification

The classification of glycemic events for each patient is displayed in figure 4. Based on this analysis, the subgroup of patients with only severe (S) and mild (m) hypo glycemic events were selected and analyzed. There were 20 patients in this group, but only 17 of them were selected for further analysis due to MA train exclusion criteria (N = 1), poor sensitivity threshold coverage across the cortex (N = 1) and missing NIRS data for SGC synchronization (N = 1 ) (N = 17 preterms, mean GA = 30 weeks, *σ* GA = ±10 days, mean BW = 1399 g, *σ* BW = ±271 g, minimum BW = 900 g and maximum BW = 1900 g). From figure 4, it can be seen that the glucose profile of the patients was highly variable with patients having only hypoglycemia (38%), only hyperglycemia (27%), both of them (33%), or none (2%), with a prevalence of mild hypoglycemic events.

**Figure 4:**
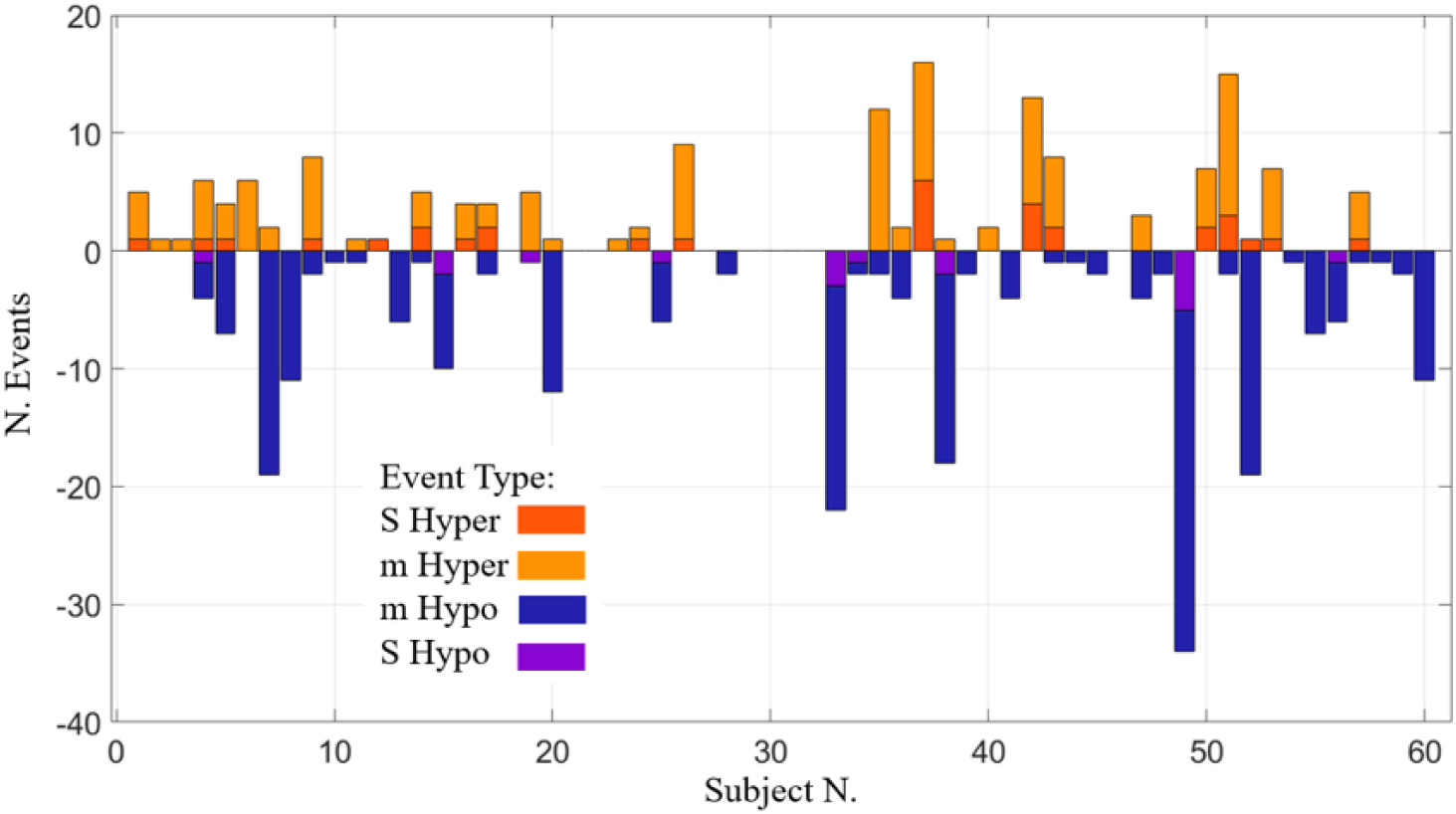
Number of glycemic event, subdivided per type (severe or mild hypo- or hyperglycemia), of all preterm patients.

### 3.2 HbT Results

In total from the 17 patients in the subgroup, summing across the three time windows, there were 372 events classified as S- and m-hypo events, 357 of these were synced with NIRS data, 236 of these passed initial signal quality checks, and 170 events passed the coverage threshold check and were used for analysis. For each time window of analysis (A, D and F), each patient could have zero, one or more glycemic events to be analyzed. Each event could result in having a different number of good measurement channels, based upon signal quality checks for that time period. This was expected due to the length of the acquisition. Figure 5 shows the average channel coverage for time window D, with the color of each measurement channel representing the number of glycemic events that channel was considered in the analysis considering all the patients and m-hypo events analyzed. Similar results were obtained for time windows A and F and for the s-hypo events (not reported).

**Figure 5:**
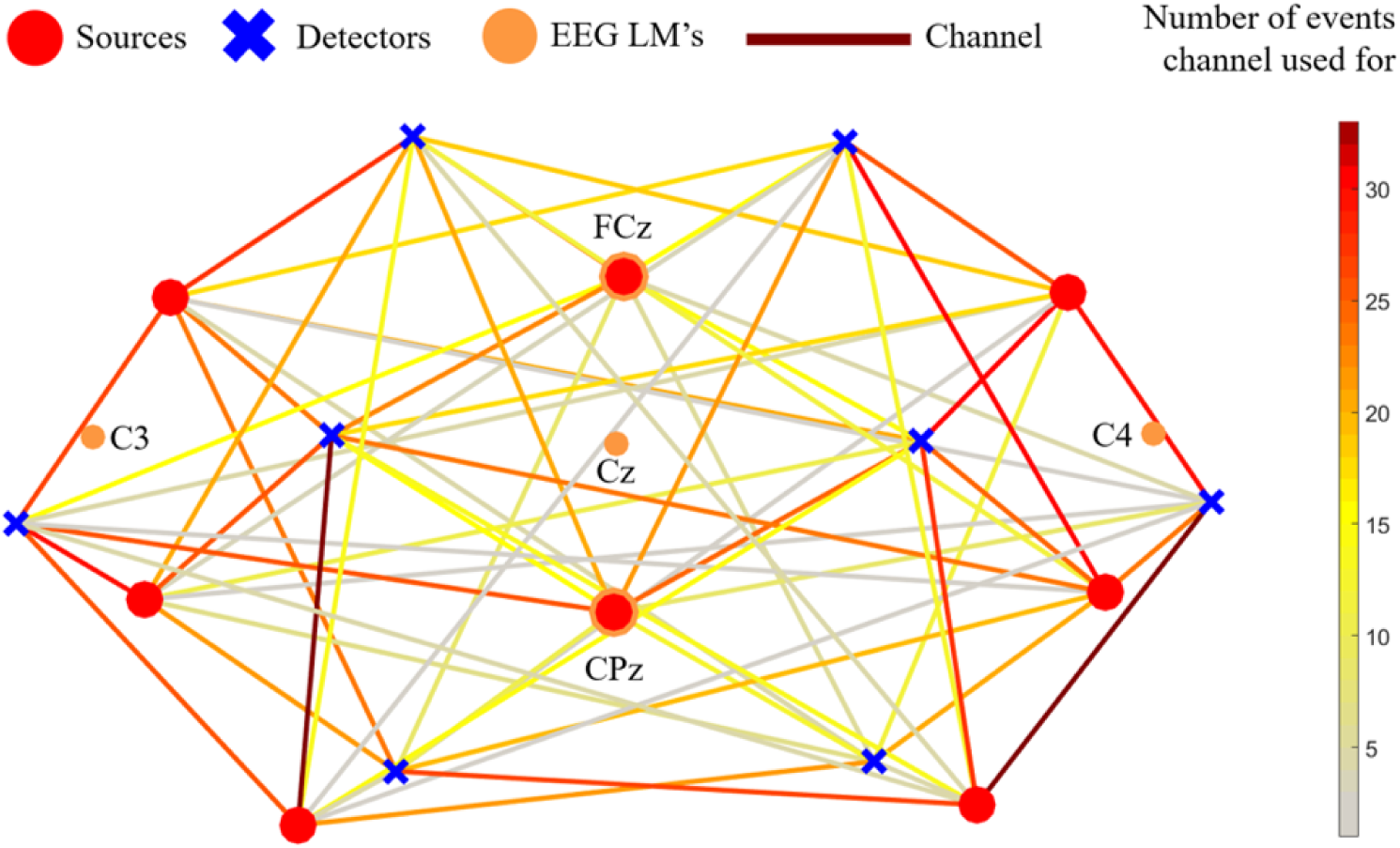
Average channel coverage. The figure displays the number of times each channel was considered for DOT analysis for all m-hypo events during time window D. For this time window, there were 50 m-hypo events from the 17 patients in the hypo-glycemic subgroup.

Figure 6 shows the net T-value for m-hypo events from all patients with at least 3 events at each time window A, D and F, as well as the net T-value for all patients within the S-m-hypo subgroup for m-hypo and S-hypo events. The spatial consistency of the events is stronger within the same patient than across all events of all the patients. However, the net T-value of all patients demonstrates that approximately half of the events seem to have a spatial coherence in some brain regions. For time window A, for example, the frontal region has a negative T-value common across many events, while the parietal area seems to be characterized by a positive net T-value. For time window D, the left frontal region has a negative net T-value across most of the patients, and the right parietal lobe has a positive net T-value across all patients. For time window F, the right motor cortex area shows a negative net T-value in most of the patients.

**Figure 6:**
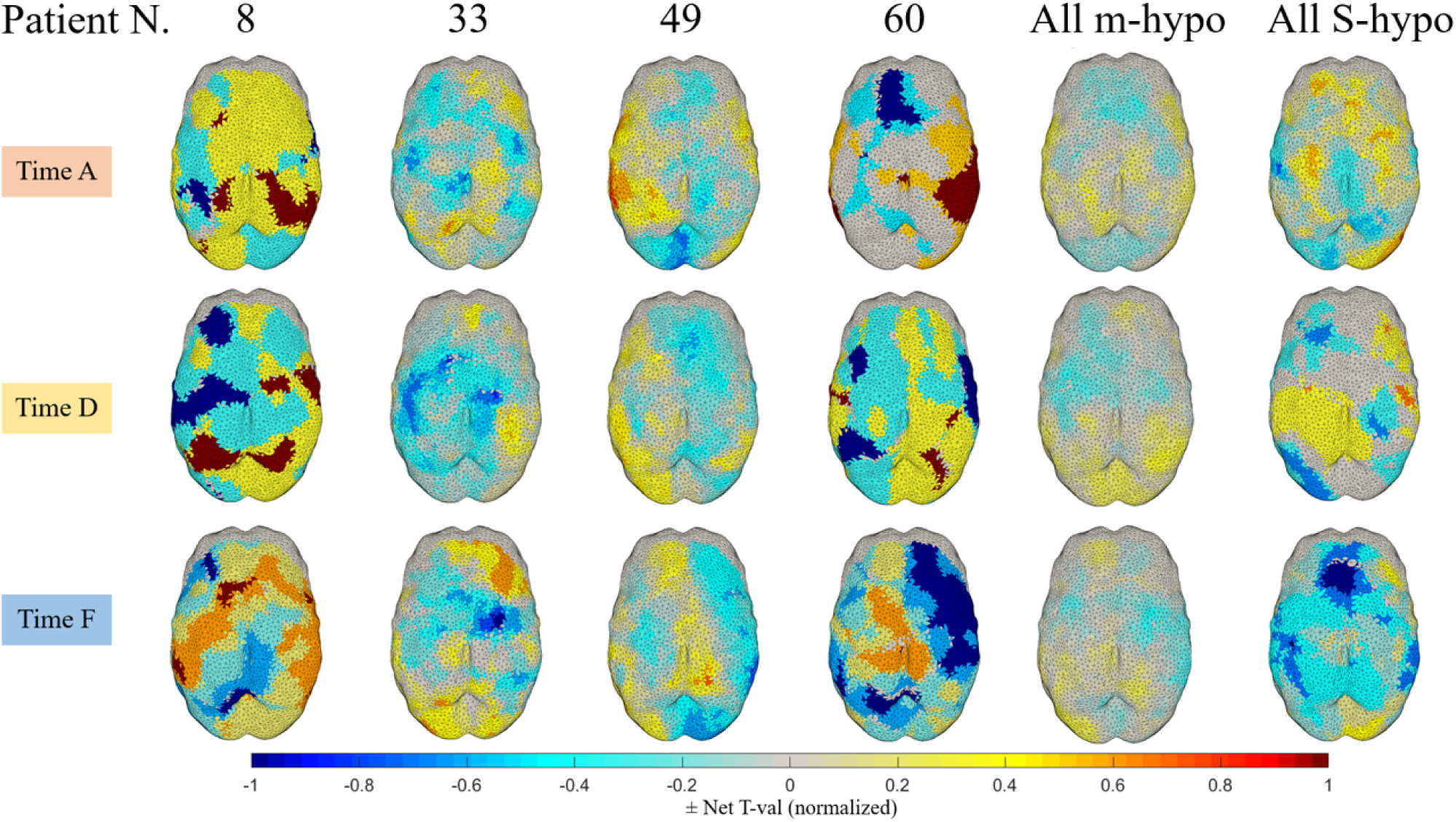
The net T-val of HbT with respect to a zero change for time windows A, D and F for patients 8 (3, 3 & 5 events, respectively), 33 (11, 13 & 10 events, respectively), 49 (14, 15 & 15 events, respectively), 60 (4, 3 & 5 events, respectively) and for all m-hypo (46, 50 & 52 events, respectively), and S-hypo (9,6 & 7 events, respectively) events of the S-m-hypo subgroup. A net T-value = 1 means that all events in that patient/subgroup had a statistically significant increase in HbT compared to zero in that location, while a net T-value = −1 means that all events in that patient/subgroup had a statistically significant decrease in HbT compared to zero in that location.

### 3.3 Correlation analyses

Figure 7 reports which HbT vs. SGC paired metrics of comparison exhibited statistically significant linear correlations (at least one cortical node with FDR corrected p-values ≤ 0.06, kept as exploratory analysis, ≤ 0.05 denoted in bold) for each time window, A, D and F and event type (m-hypo or S-hypo). A selection of these significant correlations are shown in the following, either comparing the results of the correlation using the same metrics but across different time windows or using similar metrics across the same time window. The other significant correlations are reported in supplementary Figures S1 to S18.

**Figure 7:**
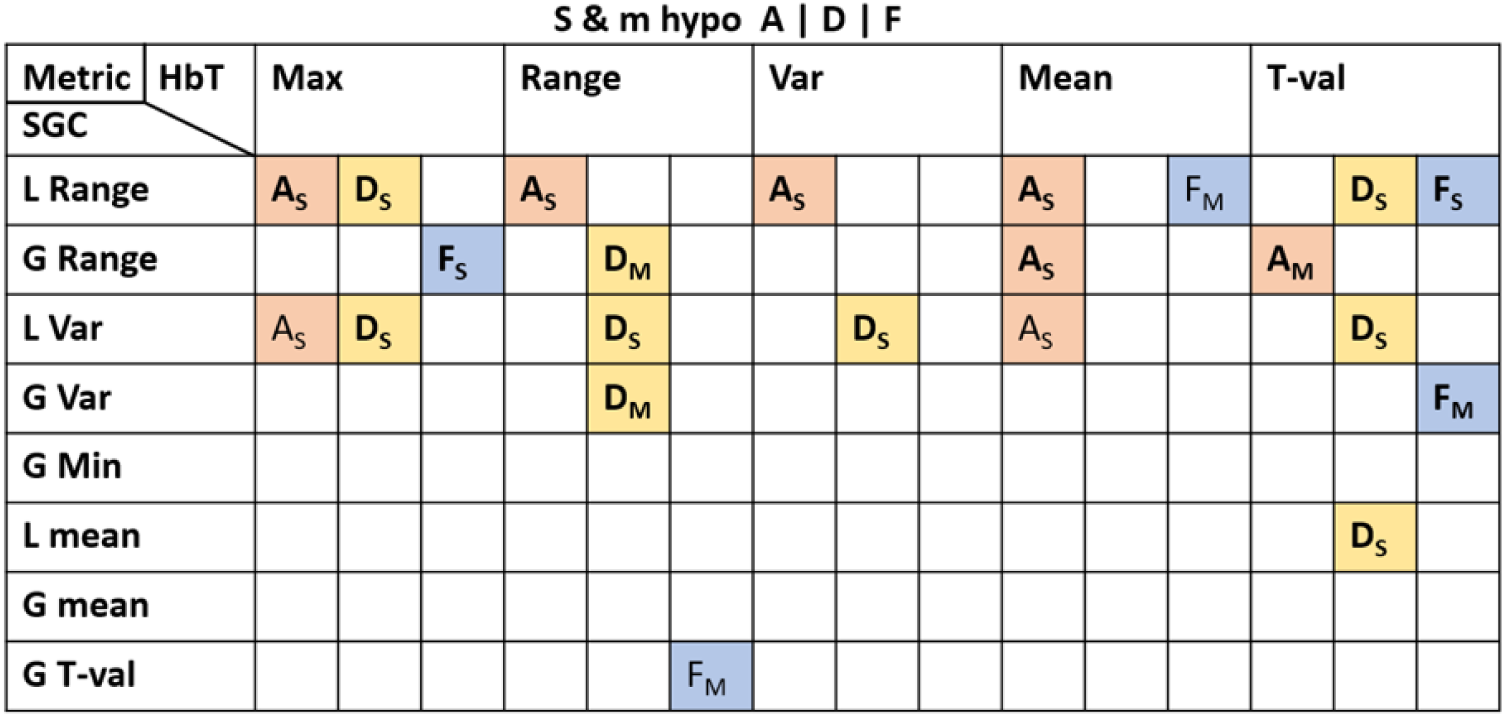
Pair of metrics that resulted statistically significant in the correlation analysis (with at least one cortical node with FDR corrected p-value ≤ 0.06, ≤ 0.05 denoted in bold). The color denotes the time window, A (peach), D (yellow) or F (blue), with the subscript denoting the type of event (M for m-hypo and S for S-hypo). For SGC metrics, L denotes ’local’ and G denotes ’Global’ changes, as described in table 4.

Figure 8 displays the results of the correlation for S-hypo events between HbT max and SGC local range for time windows A on the left and time window D on the right. The top of the figure shows the spatial R^2^ on the left and the FDR-corrected p-values on the right, while at the bottom of the figure, the correlation between the two metrics across all events is displayed for the cortical node with the highest R^2^ value. Here, the correlation results between the same metrics are different in the two time windows, showing both opposite polarity of the correlation and different brain areas involved. The only common result is a positive correlation in the left frontal areas. In most of the nodes, for time window A, smaller the SGC local range (i.e., the decrease in SGC in time window A), larger the maximum negative change in HbT, whereas for time window D, larger the SGC local range, larger the maximum positive change in HbT.

**Figure 8:**
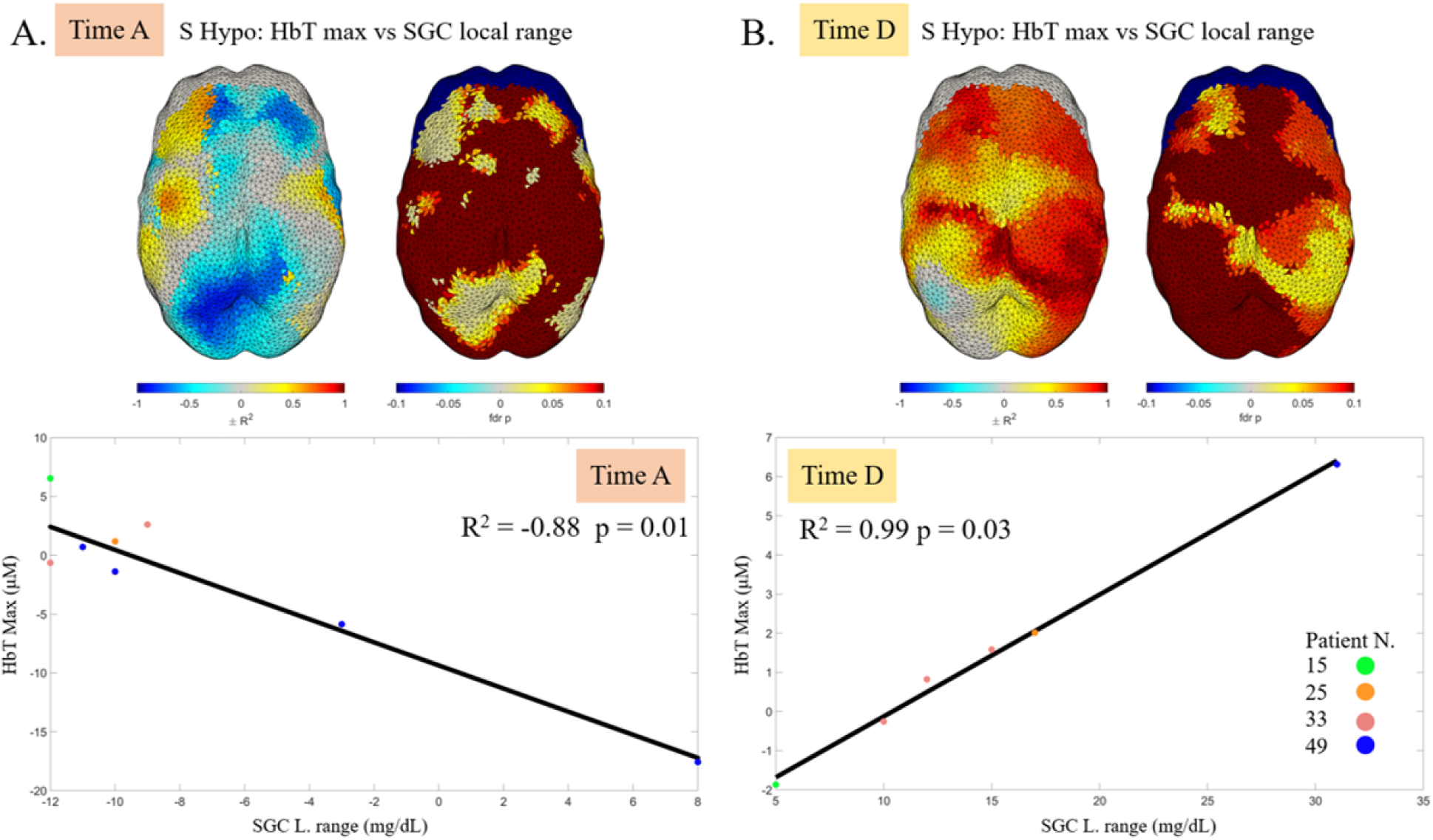
A. At the top of the figure, the R^2^ on the left and the FDR-corrected p-values on the right, for each node on the cortex displaying the correlation between HbT max and SGC local range for S-hypo events (9 events) in time window A. At the bottom of the figure is the correlation between the two metrics across all events for the cortical node with the highest R^2^ value. B. At the top of the figure, the R^2^ on the left and the FDR-corrected p-values on the right, for each node on the cortex displaying the correlation between HbT max and SGC local range for S-hypo events (6 events) in time window D. At the bottom of the figure is the correlation between the two metrics across all events for the cortical node with the highest R^2^ value. The colors of the dots, in both panels, depicts to which patient that event belonged to.

Figure 9 displays for time window D, on the left, the results of the correlation between m-hypo HbT range and SGC global range and on the right the results of the correlation between S-hypo HbT max and SGC local range. The top of the figure shows the spatial R^2^ on the left and the FDR-corrected p-values on the right, while at the bottom of the figure, the correlation between the two metrics across all events is displayed for the cortical node with the highest R^2^ value.

**Figure 9:**
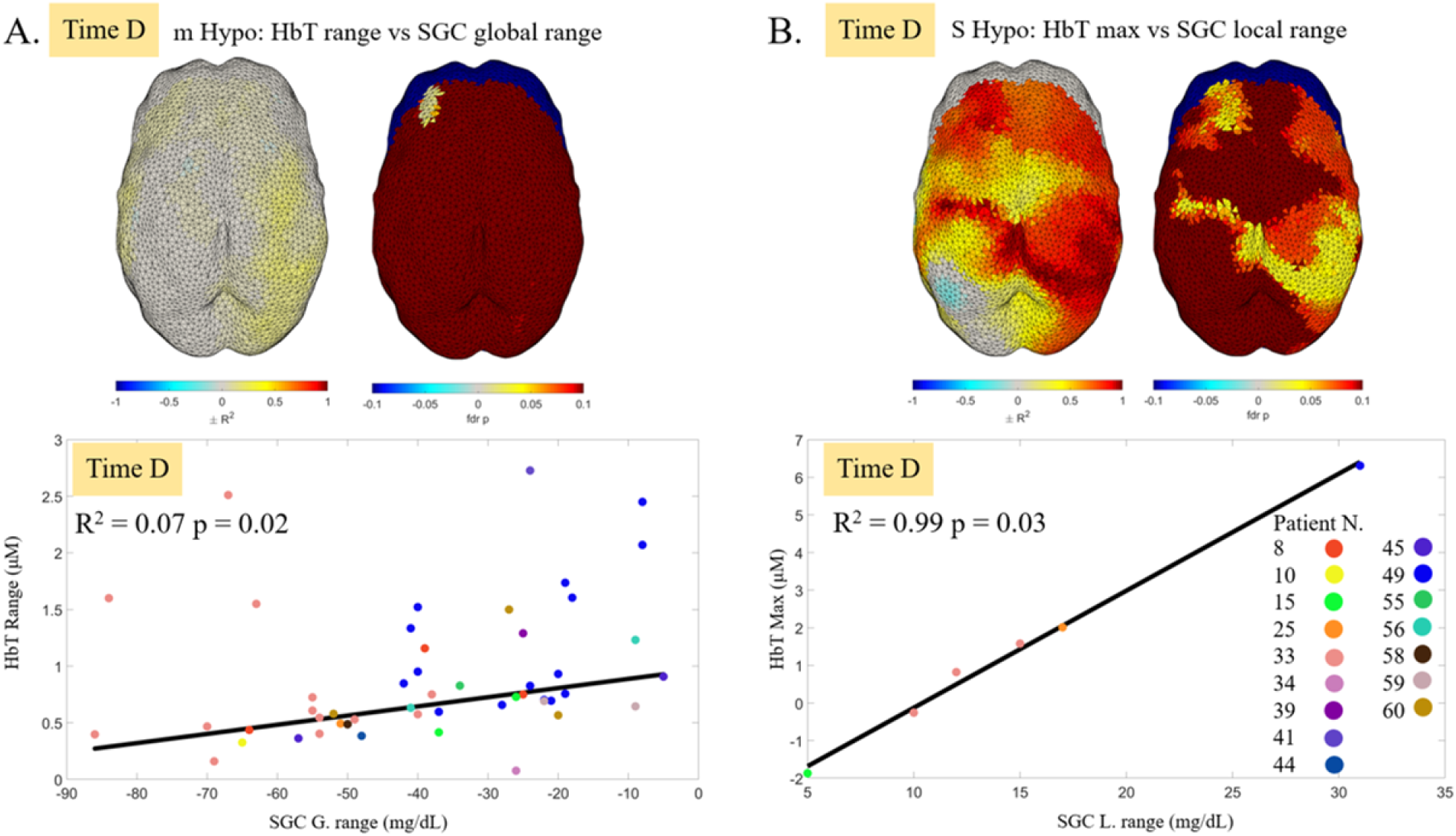
A. At the top of the figure, the R^2^ on the left and the FDR-corrected p-values on the right, for each node on the cortex displaying the correlation between HbT range and SGC global range for m-hypo events (50 events) in time window D. At the bottom of the figure is the correlation between the two metrics across all events for the cortical node with the highest R^2^ value. B. At the top of the figure, the R^2^ on the left and the FDR-corrected p-values on the right, for each node on the cortex displaying the correlation between HbT max and SGC local range for S-hypo events (6 events) in time window D. At the bottom of the figure is the correlation between the two metrics across all events for the cortical node with the highest R^2^ value. The colors of the dots, in both panels, depicts to which patient that event belonged to.

The correlation results displayed in Figure 9 have some commonalities and some differences. First, both correlations are positive and statistically significant in the left frontal lobe. There are other correlations in similar cortical areas (right parietal lobe, right motor cortex), however for m-hypo these are not statistically significant, while both do not display any correlation in the left parietal area. Second, the R^2^ values are generally higher for S-hypo compared to m-hypo and more cortical nodes survived FDR correction for the S-hypo events. Third, the interpretation of the correlation is slightly different. For m-hypo events, the larger the SGC global range (i.e., the change in SGC across the entire event from the stable euglycemic baseline), the smaller the range in HbT changes within the 10 minute period of time window D (i.e., smaller increases in blood volume). For S-hypo events, the larger the local increase in SGC in time window D, the larger the maximum HbT change occurring within the same time window.

Figure 10 displays the results of the correlation at time window F between m-hypo HbT mean and SGC local range. The left of the figure shows the spatial R^2^ and the FDR-corrected p-values, while on the right of the figure, the correlation between the two metrics across all events is displayed for the cortical node with the highest R^2^ value. These two metrics show a statistically significant positive correlation in two bilateral small areas approximately over the motor cortex. At the end of the glycemic event, in time window F, when SGC increases ⪅ 12 mg/dL, the average change in HbT is negative and stronger when there are smaller changes in SGC, whereas for SGC increases ≳ 12 mg/dL, the average change in HbT is positive and stronger when there are larger changes in SGC.

**Figure 10:**
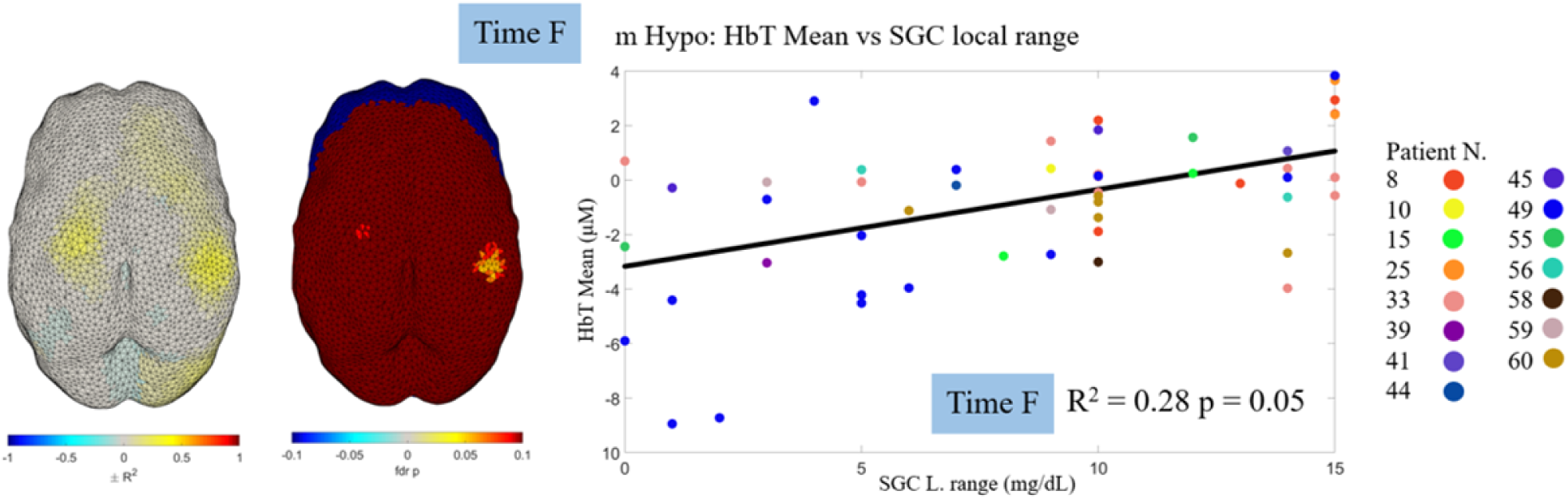
On the left of the figure, the R^2^ on the left and the FDR-corrected p-values on the right, for each node on the cortex displaying the correlation between HbT mean and SGC local range for m-hypo events (52 events) in time window F. On the right of the figure is the correlation between the two metrics across all events for the cortical node with the highest R^2^ value. It should be noted that the minimum corrected fdr p value is 0.054.

## 4 Discussion

The link between glucose concentration and cerebral hemodynamics in very preterm newborns has been investigated in previous studies (Matterberger et al., 2018, Skov and Pryds, 1992, Pryds et al., 1990), but it was not clear whether glucose variability could impact brain hemodynamics with regional specificity and how the temporal pattern of the hemodynamic response during hypo or hyperglycemia unfolds. Here, a cohort of 60 preterm newborns were recruited in the BabyGlucoLight clinical trial; newborns were monitored continuously and simultaneously with CGM and DOT for several days after birth.

The glucose profile of the patients was highly variable with patients only having hypoglycemic events, only hyperglycemic events and both types of events, approximately a third of the time each, with a prevalence of mild hypoglycemic events. This demonstrates that the glucose variability of this cohort is highly variable in the first week of life. These results suggest that preterm neonates are more prone to experiencing either hyper or hypo-glycemia, with a smaller subgroup displaying both behaviors (Angelis et al., 2023). Seventeen patients, classified as having S and/or m-hypo events only, were selected to investigate the sole impact of hypoglycemia on brain hemodynamics, reducing the number of cofounding factors between patients. DOT data were pre-processed, dealing with motion artifact detection and correction processes, and changes in HbT compared to baseline were evaluated to individuate brain regions with significant increases or decreases in cerebral blood volume in three selected time periods of each glycemic event, namely at the start and end of the event and when hypoglycemia reached its minimum value.

The spatial pattern of changes in HbT during the preselected time windows showed some variability both within and between patients but also some consistency across events and patients (figure 6). In particular, 4 patients, with at least 3 m-hypo events in each time window, were individually evaluated to investigate within-patient variability, while events of all patients, separately for mild and severe hypoglycemia, were analyzed together to investigate between-subject variability in spatial pattern. Results showed a higher spatial consistency for S-hypo than m-hypo events. One explanation could be the higher number of m-hypo events in the dataset, which likely increased the variability. However, these results could also indicate that the spatial pattern of the hemodynamic response is more consistent across patients and events when responding to a stronger decline in glucose concentration. The categorization of glucose events into mild and severe is based on standardized operational thresholds, but there is still little evidence for what the actual optimal individual target glucose levels at birth should be (Hay et al., 2009). In this perspective, it is highly likely that several m-hypo events, above all those with a minimum glucose concentration very near the operational threshold for mild hypoglycemia, could have actually been normal glucose fluctuations for that individual patient. This hypothesis could explain the lower spatial consistency observed for m-hypo events. Looking at both the individual patients and the entire dataset of events, some cortical areas seem to show higher spatial consistency than others, which might indicate a more common and reactive hemodynamic response in these areas to changes in glucose concentration. For example, when hypoglycemia reached its minimum, the left frontal and motor areas exhibited a decrease in HbT across all m-hypo events and also in the individual patients; this pattern was similar for S-hypo events, although the decrease was more localized in the frontal areas. In the same time window, for all individual patients and all m-hypo events, the right parietal area showed, instead, an increase in HbT, while in the same area a decrease in HbT was obtained during S-hypo events. At the start of hypoglycemia, the central frontal area exhibited a decrease in HbT across all m-hypo events and also in the individual patients (apart from patient 8), while in the same area an increase in HbT was obtained during S-hypo events. At the end of hypoglycemia, the left frontal area showed an increase in HbT, while in the same area a decrease in HbT was obtained during S-hypo events. In the same time window, for both mild and severe hypoglycemia, the right motor cortex exhibited a decrease in HbT. These results suggest not only that the brain hemodynamic response to glucose changes is not global but varies locally, highlighting the importance of monitoring the whole brain, but that the brain hemodynamic response might differ in certain cortical areas depending on the severity of the glucose event.

Previous MRI studies reported that occipital, but also parietal areas, are the most susceptible to neonatal hypoglycemia (Zhang et al., 2022, Tam et al., 2008,Filan et al., 2006, Burns et al., 2008, Barkovich et al., 1998 and Alkalay et al., 2005), although it has been reported that, compared to full-term newborns, preterm were less affected by hypoglycemia, showing no brain damage in these areas (Caksen et al., 2011). In this study, we could not monitor occipital areas, since the optodes would have been uncomfortable for the newborns due to the long acquisition, but only parietal ones. An MRI-discovered injury in a cortical area might be due to a previous lack of oxygen during hypoglycemia, following a missing compensatory increase in blood flow (Skov and Pryds, 1992). At the minimum of the S-hypo events, the area with the most common decrease in HbT is the left parietal area, followed by the left frontal area, which could indicate that these areas might be more susceptible to brain injury than other areas, where other compensatory effects might occur. The left fronto-parietal network and the left intraparietal sulcus (IPS) are key cortical areas for several cognitive processes, such as memory and attention (Cowan et al., 2011 and Scolari et al., 2015), the precursor of executive functions, which were found to be impaired during development in preterm newborns suffering from hypoglycemia at birth (Shah et al., 2019).

To investigate whether specific brain areas are more susceptible to specific features of the change in glucose concentration, as well as its severity, several correlations were computed between HbT and SGC data. Five metrics were derived from the HbT data and 8 metrics from the SGC data, giving a total of 40 comparisons each across the 3 time windows, with 22 of these comparisons showing statistically significant correlations after correction for multiple comparison (4 of these 22 had a p values ≤ 0.06 and were kept as exploratory; figure 7). In particular, the highest number of correlations were found for S-hypo events (16/22) in time windows A (7/16) and D (7/16). M-hypo events had fewer significant correlations, distributed in the three time windows. These results might indicate that the start of the events and the minima of the events could be more important for the coupling between glucose and brain hemodynamics, or they might simply be more common and generalizable among patients and events, being the moment when glucose starts to decrease significantly and the time when glucose reaches its minimum value. Results in time window F might be more impacted and influenced by the duration of the hypoglycemia, which is very different in this population (maximum length = 495 minutes and minimum length = 55 minutes). Furthermore, these results seem to be in agreement with results reported in Fig. 6, given the higher number of correlations reported for S-hypo than m-hypo events. Concerning the analyzed metrics, furthermore, three SGC metrics resulted in driving the correlation with HbT: the local range, the local variance and the global range. The local and global SGC mean metrics could be influenced by the severity and stability of the glucose values within the time windows and the duration of the euglycemic baselines, respectively, therefore being a multi-factorial metric, which was hardly directly correlated with brain hemodynamics. The global minimum was never correlated with brain hemodynamics in any of the time windows, likely indicating that the brain response to a specific glucose value might be more individualized and less generalizable. This is in line with the idea that operational thresholds are not ideal and that optimal individual target glucose levels at birth are still unknown (Hay et al., 2009). The global T-val was hardly directly correlated with brain hemodynamics as well; the t-val is dependent on the length of the hypoglycemia and its severity and it might be that brain hemodynamics is not responding linearly to these two features.

In the following paragraphs, we discuss the most significant correlations (those with enough fdr-corrected significant nodes to create small ROIs) obtained between brain hemodynamics and the main three SGC metrics.

The correlations between the maximum, the range, the variance and the mean of HbT and SGC local range in time window A (start of the event) for S-hypo events yielded similar results (Figure 8 and supplementary figures 1-3), with a strong negative correlation in the left parietal and right frontal areas and a weaker positive correlation in the left frontal and right parietal areas. These results suggest that the left parietal and right frontal areas respond with a compensatory increase in HbT: the larger the decrease of SGC, the larger the increase in HbT. These results agree with the hypothesis of the existence of a cerebral glucose sensor, whereby when starting hypoglycemia, the preterm brain reacts by vasodilatation of the capillaries to increase the cerebral blood flow and therefore the glucose flow to the brain (Skov and Pryds, 1992). However, our results also show that in the right parietal and left frontal areas this pattern is inverted and the other cortical areas do not appear to be susceptible to glucose changes, therefore highlighting the spatial specificity of the cortical hemodynamic response to the start of severe hypoglycemia.

The correlation between the maximum of HbT and SGC local range in time window D (minimum of the event) for S-hypo events showed a different pattern compared to the same correlation in time window A (fig. 8B). Similar results, although with an extended cortical area of significant nodes, were found correlating the maximum, range and variance of HbT with the local SGC variance. Here, a strong positive correlation was found in the bilateral motor area and in the right parietal one. Similarly to the start of the events, a positive correlation was observed also in the left frontal area. These results suggest that, when in severe hypoglycemia, these areas of the brain respond with a stronger increase in HbT, relative to the HbT level immediately before reaching the minimum of hypoglycemia, when the glucose level starts to go back toward euglycemia with a steeper change (large positive local range). This might be due to the effect of nutrients and/or glucose boluses infused to contrast hypoglycemia, which increased glucose availability and led to the resolution of the hypoglycemic event. In contrast, when still in severe hypoglycemia (small positive local range), brain hemodynamics might be in a steady state, or saturated state, showing no compensatory increases nor decreases of HbT during the hypoglycemic minimum. Interestingly, in time window D the left parietal area, which showed the strongest correlation in time window A, did not show any correlation between brain hemodynamics and SGC.

The correlation between the range of HbT and SGC global range in time window D (minimum of the event) for m-hypo events showed a similar, although weaker, pattern as the one obtained for the S-hypo events in the same time window (fig. 9A), with a positive correlation in the left frontal and right parietal areas. Here, however, the interpretation of these results is slightly different. These results suggest that at the minimum of mild hypoglycemia, brain hemodynamics showed larger changes (independently of the sign of the change) for milder events (small SGC range) and smaller changes for stronger changes in glucose concentration. This latter could indicate that, similarly to severe hypoglycemic events, also for mild hypoglycemic events, brain hemodynamics is in a steady saturated state when hypoglycemia is more severe. For milder events, instead, brain hemodynamics is still changing, either trying to compensate for the lack of glucose or showing a decrease in CBV in these areas.

The other significant correlation for m-hypo events was the one between the T-val of HbT and the global SGC range in time window A (supplementary figure 7). Here, a small region in the left frontal area exhibited a positive correlation, pointing to an initial compensatory response of brain hemodynamics (positive T-val) for milder hypoglycemic events and an immediate decrease in HbT for stronger glucose changes. Most of the cortex, however, did not show any correlation between these two metrics.

The end of the glucose event (time window F) displayed the lowest number of significant correlations, both for severe and mild events. The only meaningful correlations here are the one between HbT mean and local SGC range for mild events (figure 10) and the one between the T-val of HbT and local SGC range for severe events (supplementary figure 15). For mild events, two bilateral spots in the motor cortex displayed a positive correlation, with stronger negative HbT mean values for smaller changes in local SGC range, while for severe events, a few spots mostly in the right hemisphere showed a negative correlation, with a steeper return to euglycemia characterized by a more negative change in HbT. These results are hard to interpret, likely because, as already stated, brain hemodynamics in time window F might be more susceptible to the duration of the event and the pattern of return to euglycemia, which were not investigated in this analysis. HbT changes are considered vs. the 2 minutes of baseline preceding the return to euglycemia and therefore, without analyzing the speed of glucose changes after the minimum of hypoglycemia, their interpretation might be biased. Indeed, the threshold selection of glycemic events, rate of glucose change and events duration may all play toward the measured hemodynamic changes, however with this approach, we were not able to quantify their impact. Future works should consider this feature in the analysis of brain hemodynamic behavior at the end of a hypoglycemic event. There are no previous works to directly compare our results at the end of the events with, since previous aforementioned studies have not investigated cerebral hemodynamics at the end of hypoglycemia. However, one study found that approximately 30 minutes after intravenous glucose injection in hypoglycemic neonates, CBF decreased by an average of 11.3% (Pryds et al., 1990). Results in time window F might be displaying that a reduction in cerebral blood volume is still ongoing when the hypoglycemic events comes back to euglycemia and likely requires some further minutes to stabilize.

This study has a few limitations. The first main limitation of the study is that the measurements of HbT are relative changes in concentration with respect to a baseline, and, due to the length of the acquisition and the strong presence of motion artifacts, a different baseline (i.e., the 2 minutes before each considered feature of the glucose curve, namely, the start, end and minimum) had to be used. Therefore, the metrics derived from HbT data do not take into account the absolute value of HbT, which may provide more relevant information to compare with the absolute values of SGC. This was partly mitigated by deriving metrics of SGC both at the local and global level, to try to characterize hemodynamic changes in relation to both the concurrent glucose changes and the whole glucose profile. A future study could use time domain NIRS/DOT, which can measure absolute concentrations of HbT (Calcaterra et al., 2024), instead of continuous wave NIRS/DOT, which is limited to only measuring changes in HbT.

Another limitation of this study is the lack of information on individual optode positions, since it was not possible to digitize the positions of the cap on the preterm’s head. Furthermore, for comparison purposes, all reconstructions were performed on the 30-week GA head model, instead of their age specific model. These two issues might have increased the reconstruction error. It is also worth noting that, due to the nature of the experiment, with continuous acquisitions of multiple days of duration during which standard clinical procedures were performed on the newborn, a robust MA detection and correction processing pipeline had to be implemented. Given the number of patients and glycemic events, a variety of MAs were observed in the data, which all had to be processed automatically. Because of this, there may have been cases where a specific MA was not corrected optimally, or cases for given events where MAs were over or under classified. However, the methods for MA detection (Sherafati et al., 2020 and Huppert et al., 2009) and correction (Yang et al., 2022 and Di Lorenzo et al., 2019) employed here have been extensively tested in the literature. Therefore, they should be adequate for a generalized approach.

## 5 Conclusion

This study reports the first results of the BabyGlucoLight trial, during which preterm newborns were continuously and simultaneously monitored with DOT and CGM. Only preterms with hypoglycemic events were selected and analyzed in three selected time windows of the hypoglycemic event, namely the start, the end, and the minimum of hypoglycemia. Changes in HbT in these three time windows showed spatial consistency across patients and events, above all for the severe hypoglycemic events. The left fronto-parietal network, a key area for attention and memory processes, consistently showed a decrease in HbT at the minimum of hypoglycemia, suggesting that these areas might be suffering more than others. Several correlations between metrics extracted from the HbT changes and SGC were obtained, showing specific spatial patterns for different time windows and depending on the severity of the event. In particular, a higher number of correlations was obtained for severe than mild hypoglycemic events. These results suggest that not only changes in HbT and SGC are linked during hypoglycaemia, but that different areas of the brain respond differently depending on the specific hypoglycaemic event. In conclusion, HbT changes in response to severe hypoglycemic events seem to be more spatially consistent and more correlated with SGC than mild events. Further results of the BabyGlucoLight trial will evaluate how this coupling affects neurodevelopment, as well as investigate whether hyperglycaemia shows similar coupling and spatial correlations, and if tighter glucose control could lead to a better outcomes for the preterm.

## Data and Code Availability

The code used for this study can be found on Github at https://github.com/sbrigadoi/BabyGlucoLight. The data for this study can be found at https://researchdata.cab.unipd.it/id/eprint/1518.

## Author Contributions

The contributions are denoted using CRediT (Contributor Roles Taxonomy) terminology, whereby the initials of the authors are used, conceptualization GP, SG, GR, FS, DT, EP, AG, EB, SB, methodology GP, AG, SB, investigation GR, FS, DT, EP, SG, software GP, formal analysis GP, SB, writing (original draft) GP, SB, writing (review & editing) SG, GR, FS, DT, EP, AG, EB and funding acquisition AG, EP, SB.

## Supplementary Material

The supplementary material shows all cortex graphs of statistically significant correlations between HbT and SGC metrics and can be found in the accompanying document ”Supplementary Figures Word Doc.docx”.

