## Supplementary Figures for "Investigating brain haemodynamics during hypoglycaemia in very preterm neonates using diffuse optical tomography"

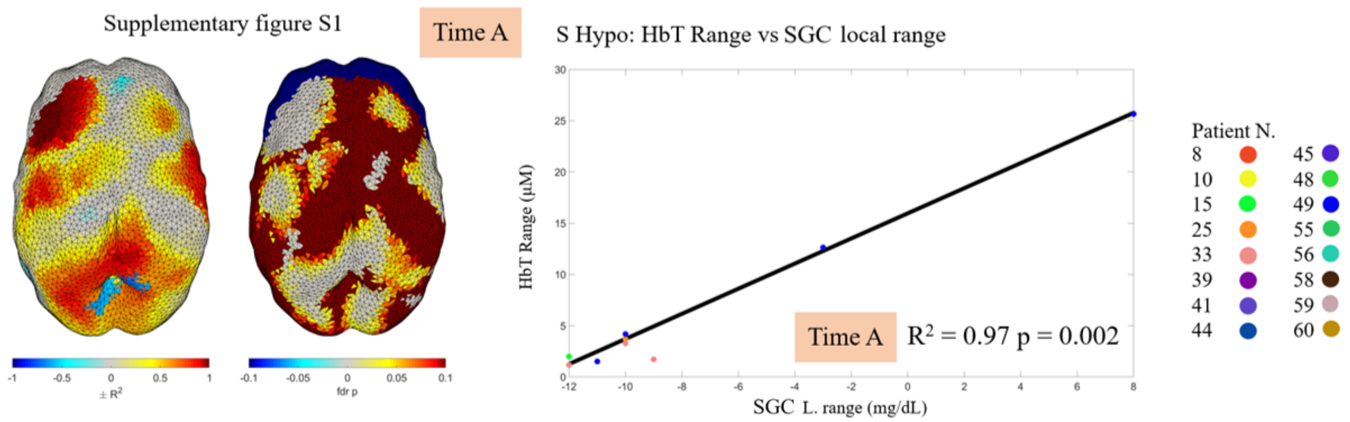


S1: On the left of the figure, the $R^2$ on the left and the FDR-corrected p-values on the right, for each node on the cortex displaying the correlation between HbT range and SGC local range for S-hypo events (9 events) in time window A. At the bottom of the figure is the correlation between the two metrics across all events for the cortical node with the highest $R^2$ value.

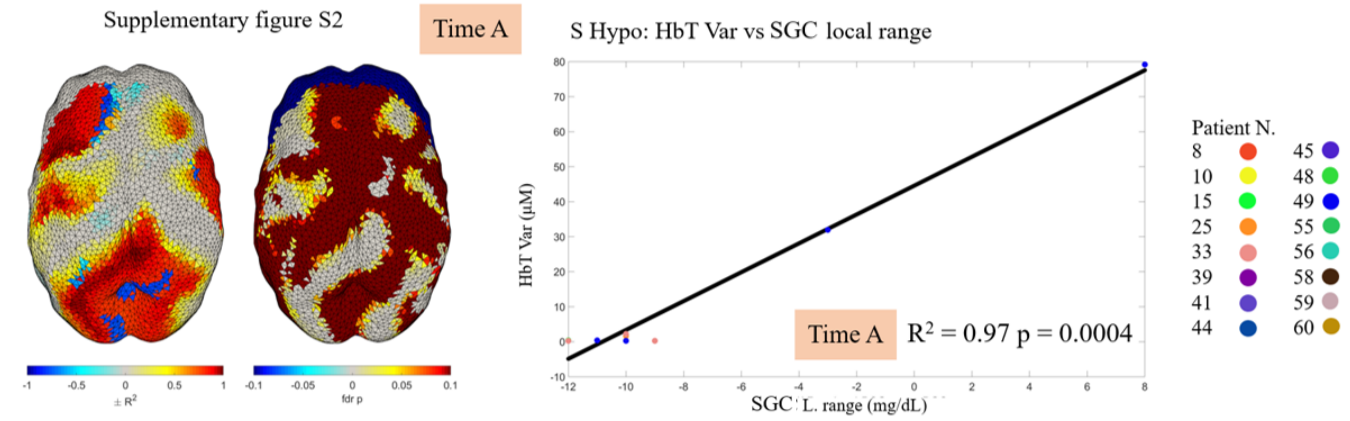


S2: On the left of the figure, the $R^2$ on the left and the FDR-corrected p-values on the right, for each node on the cortex displaying the correlation between HbT var and SGC local range for S-hypo events (9 events) in time window A. At the bottom of the figure is the correlation between the two metrics across all events for the cortical node with the highest $R^2$ value.

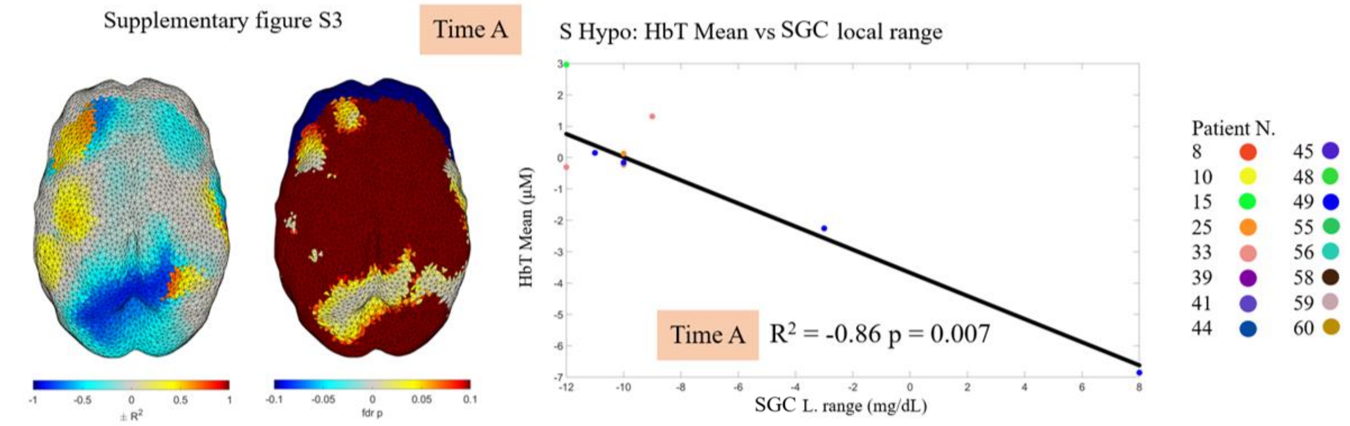


S3: On the left of the figure, the $R^2$ on the left and the FDR-corrected p-values on the right, for each node on the cortex displaying the correlation between HbT and SGC local range for S-hypo events (9 events) in time window A. At the bottom of the figure is the correlation between the two metrics across all events for the cortical node with the highest $R^2$ value.

Supplementary figure S4

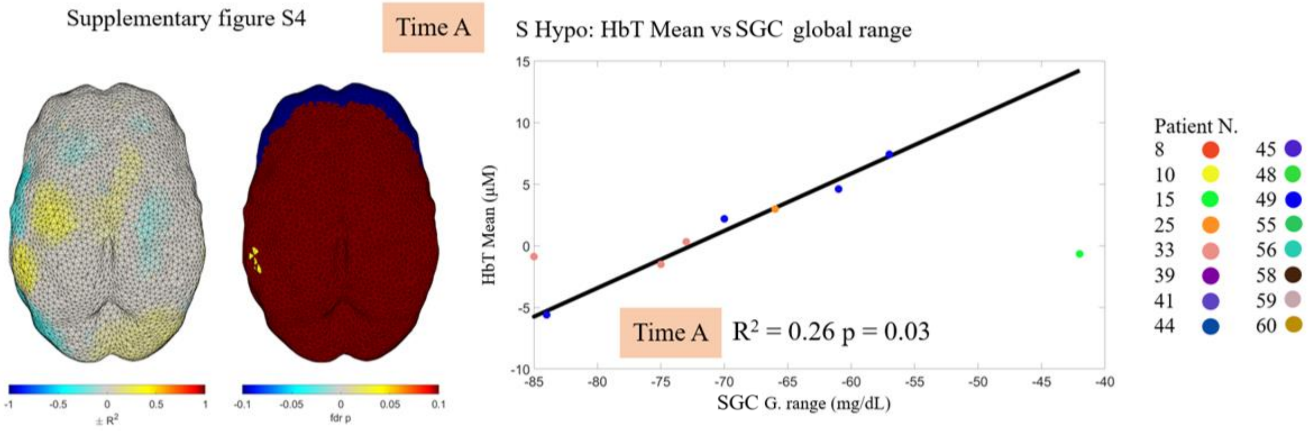


S4: On the left of the figure, the $R^2$ on the left and the FDR-corrected p-values on the right, for each node on the cortex displaying the correlation between HbT mean and SGC global range for S-hypo events (9 events) in time window A. At the bottom of the figure is the correlation between the two metrics across all events for the cortical node with the highest $R^2$ value.

Supplementary figure S5

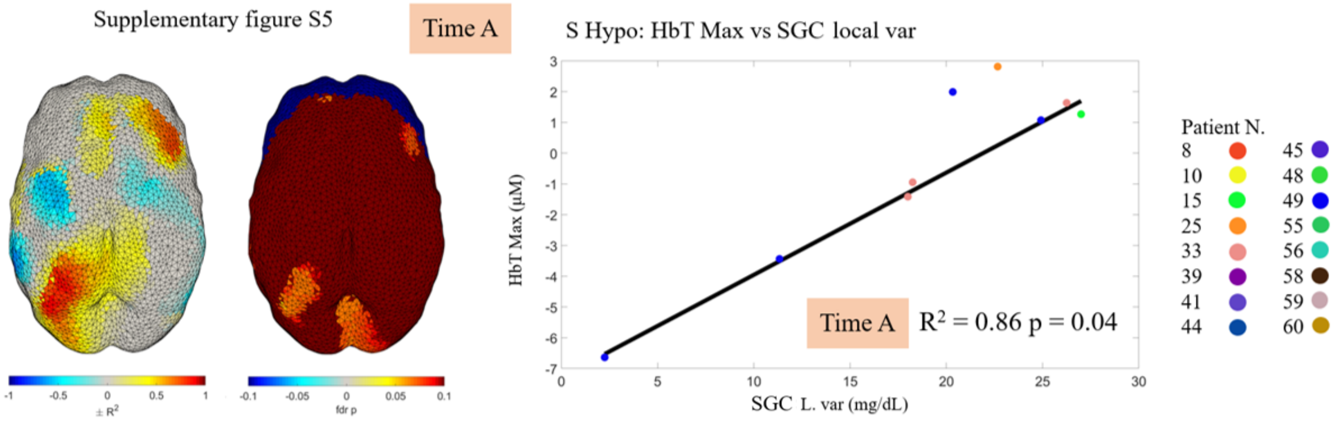


S5: On the left of the figure, the $R^2$ on the left and the FDR-corrected p-values on the right, for each node on the cortex displaying the correlation between HbT max and SGC global range for S-hypo events (9 events) in time window A. At the bottom of the figure is the correlation between the two metrics across all events for the cortical node with the highest $R^2$ value.

Supplementary figure S6

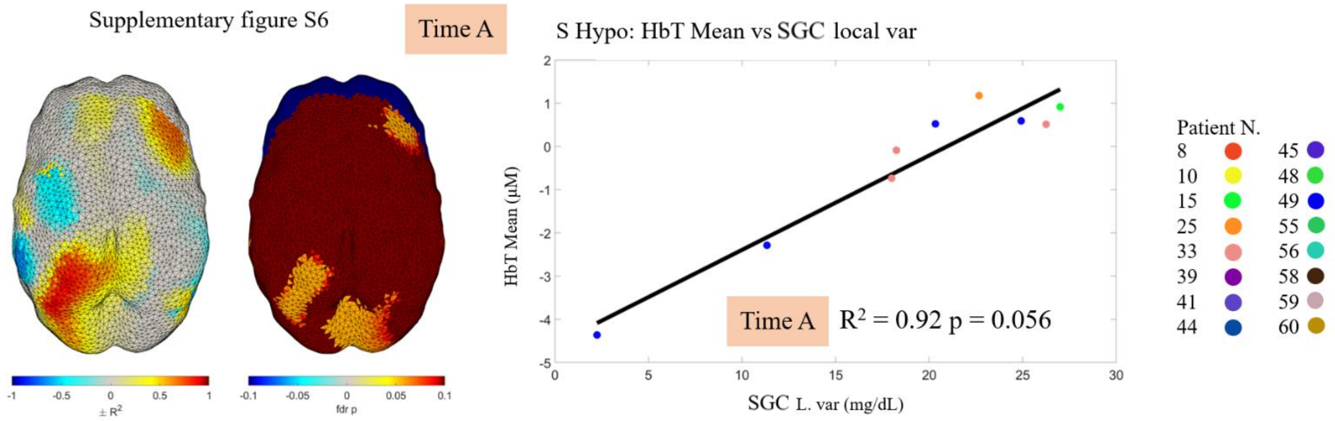


S6: On the left of the figure, the $R^2$ on the left and the FDR-corrected p-values on the right, for each node on the cortex displaying the correlation between HbT mean and SGC local var for S-hypo events (9 events) in time window A. At the bottom of the figure is the correlation between the two metrics across all events for the cortical node with the highest $R^2$ value.

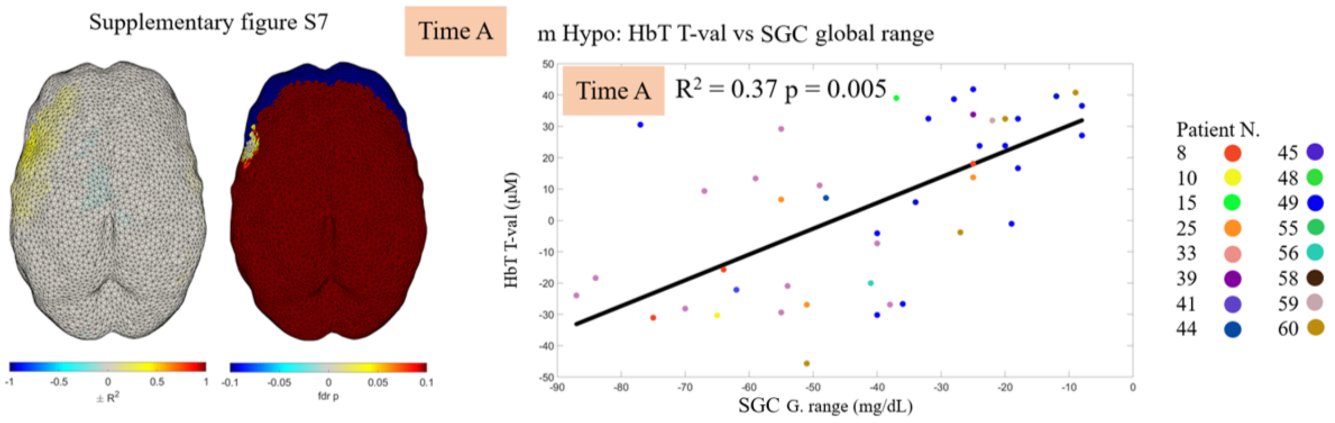


S7: On the left of the figure, the $R^2$ on the left and the FDR-corrected p-values on the right, for each node on the cortex displaying the correlation between HbT T-val and SGC global range for m-hypo events (46 events) in time window A. At the bottom of the figure is the correlation between the two metrics across all events for the cortical node with the highest $R^2$ value.

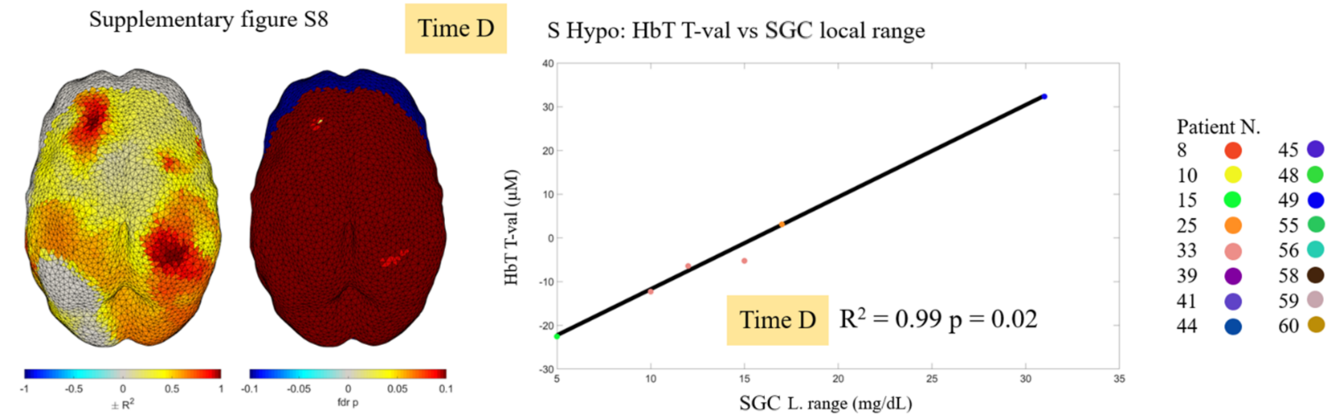


S8: On the left of the figure, the $R^2$ on the left and the FDR-corrected p-values on the right, for each node on the cortex displaying the correlation between HbT T-val and SGC local range for S-hypo events (6 events) in time window D. At the bottom of the figure is the correlation between the two metrics across all events for the cortical node with the highest $R^2$ value.

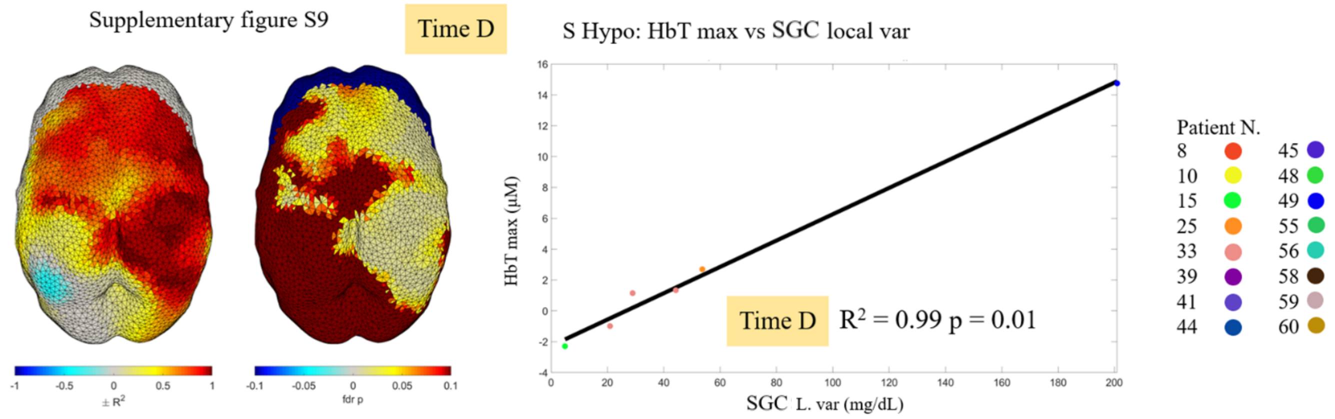


S9: On the left of the figure, the $R^2$ on the left and the FDR-corrected p-values on the right, for each node on the cortex displaying the correlation between HbT max and SGC local var for S-hypo events (6 events) in time window D. At the bottom of the figure is the correlation between the two metrics across all events for the cortical node with the highest $R^2$ value.

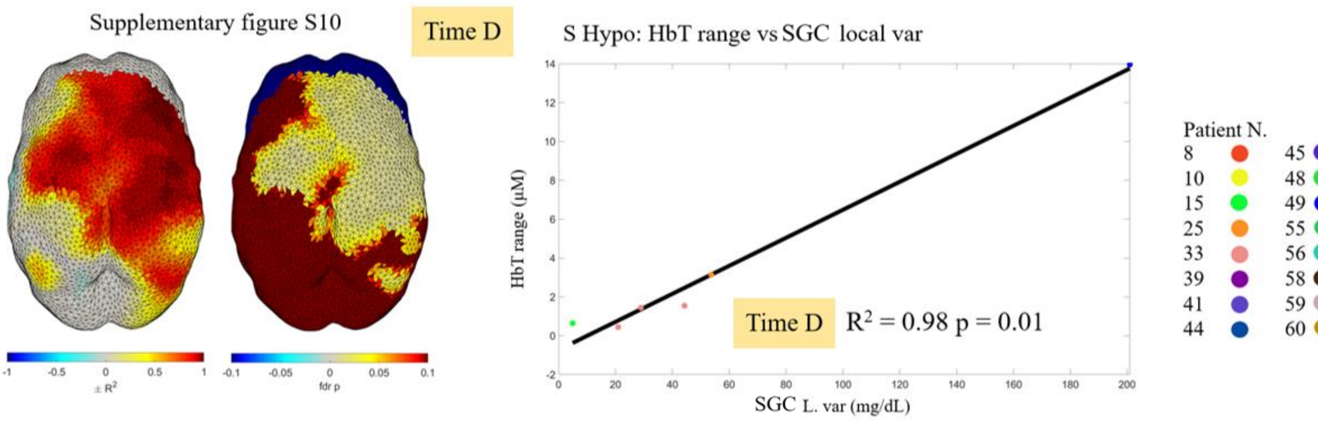


S10: On the left of the figure, the $R^2$ on the left and the FDR-corrected p-values on the right, for each node on the cortex displaying the correlation between HbT range and SGC local var for S-hypo events (6 events) in time window D. At the bottom of the figure is the correlation between the two metrics across all events for the cortical node with the highest $R^2$ value.

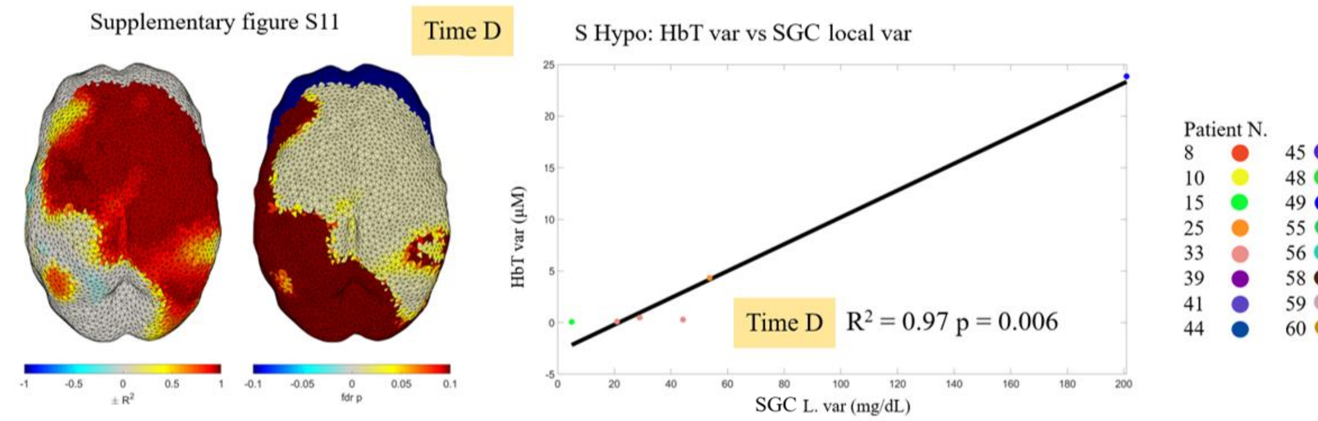


S11: On the left of the figure, the $R^2$ on the left and the FDR-corrected p-values on the right, for each node on the cortex displaying the correlation between HbT var and SGC local var for S-hypo events (6 events) in time window D. At the bottom of the figure is the correlation between the two metrics across all events for the cortical node with the highest $R^2$ value.

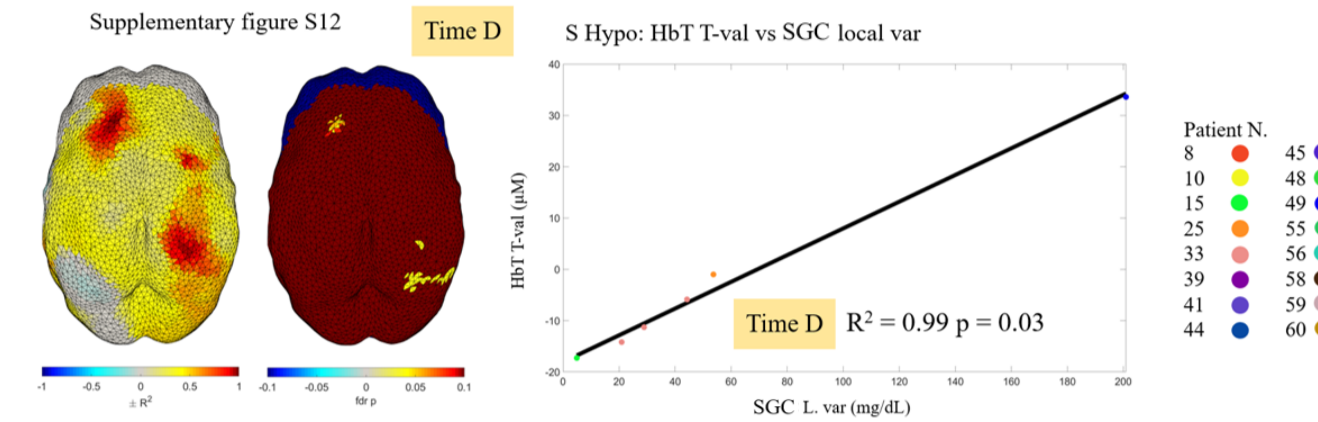


S12: On the left of the figure, the $R^2$ on the left and the FDR-corrected p-values on the right, for each node on the cortex displaying the correlation between HbT T-val and SGC local var for S-hypo events (6 events) in time window D. At the bottom of the figure is the correlation between the two metrics across all events for the cortical node with the highest $R^2$ value.

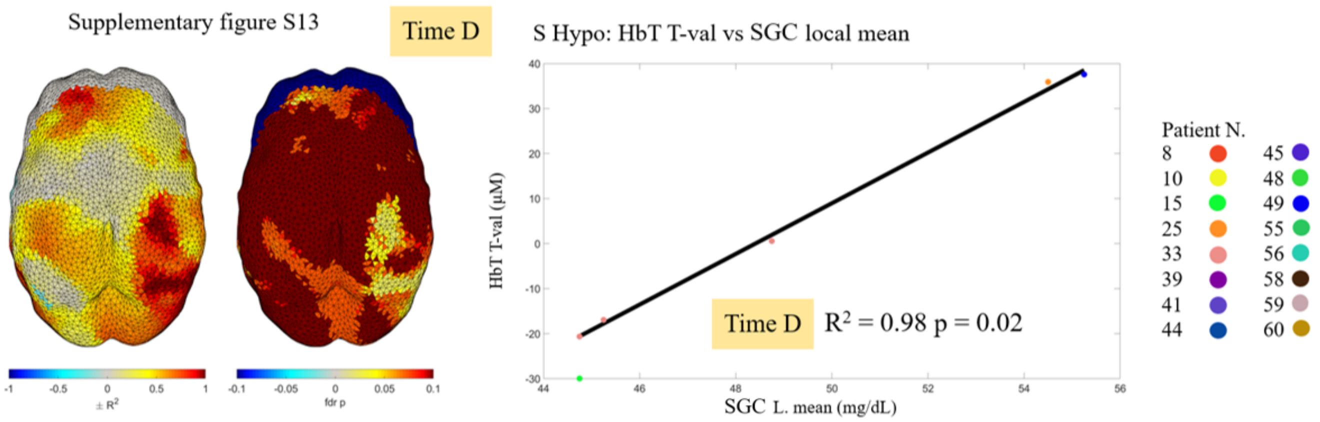


S13: On the left of the figure, the $R^2$ on the left and the FDR-corrected p-values on the right, for each node on the cortex displaying the correlation between HbT T-val and SGC local mean for S-hypo events (6 events) in time window D. At the bottom of the figure is the correlation between the two metrics across all events for the cortical node with the highest $R^2$ value.

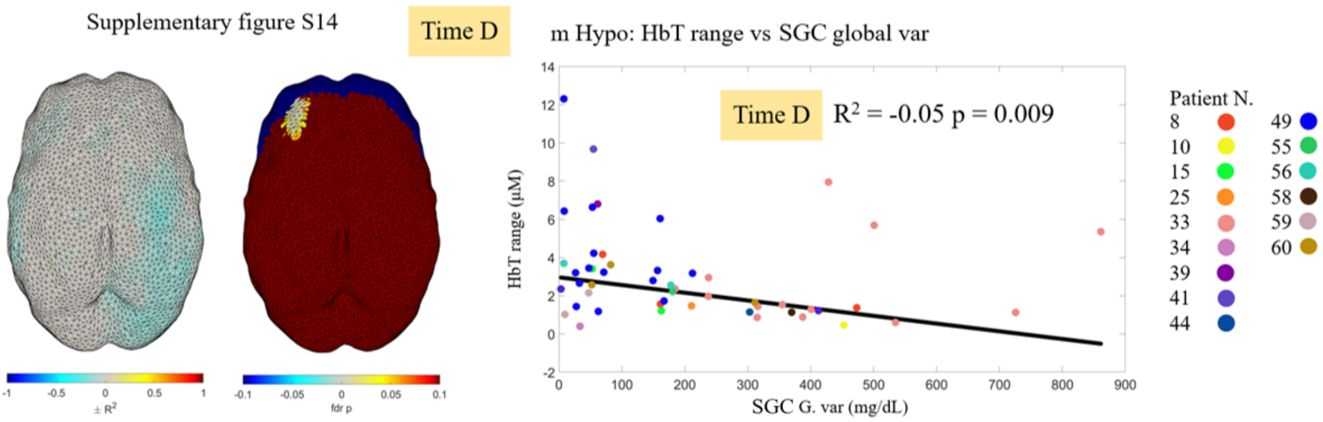


S14: On the left of the figure, the $R^2$ on the left and the FDR-corrected p-values on the right, for each node on the cortex displaying the correlation between HbT range and SGC global var for m-hypo events (50 events) in time window D. At the bottom of the figure is the correlation between the two metrics across all events for the cortical node with the highest $R^2$ value.

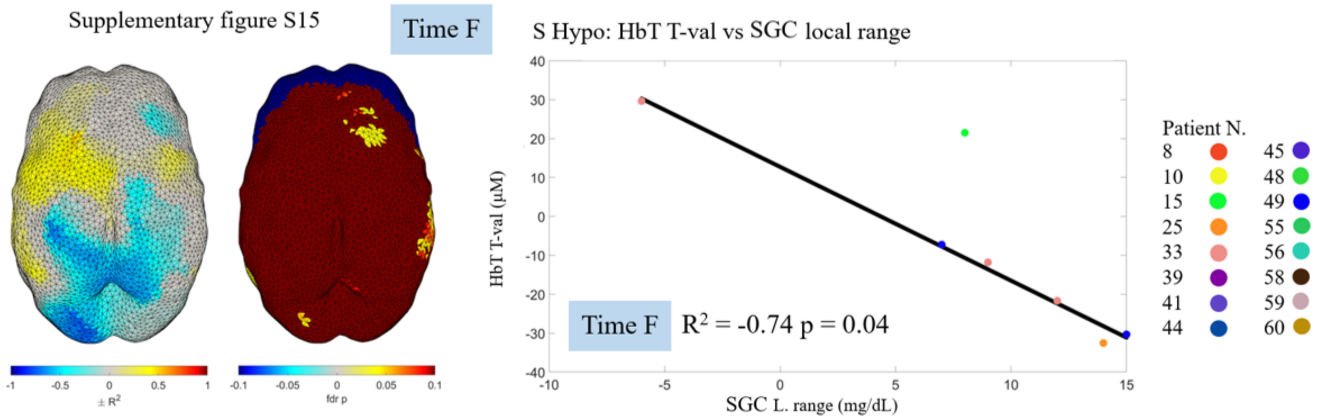


S15: On the left of the figure, the $R^2$ on the left and the FDR-corrected p-values on the right, for each node on the cortex displaying the correlation between HbT T-val and SGC local range for S-hypo events (7 events) in time window F. At the bottom of the figure is the correlation between the two metrics across all events for the cortical node with the highest $R^2$ value.

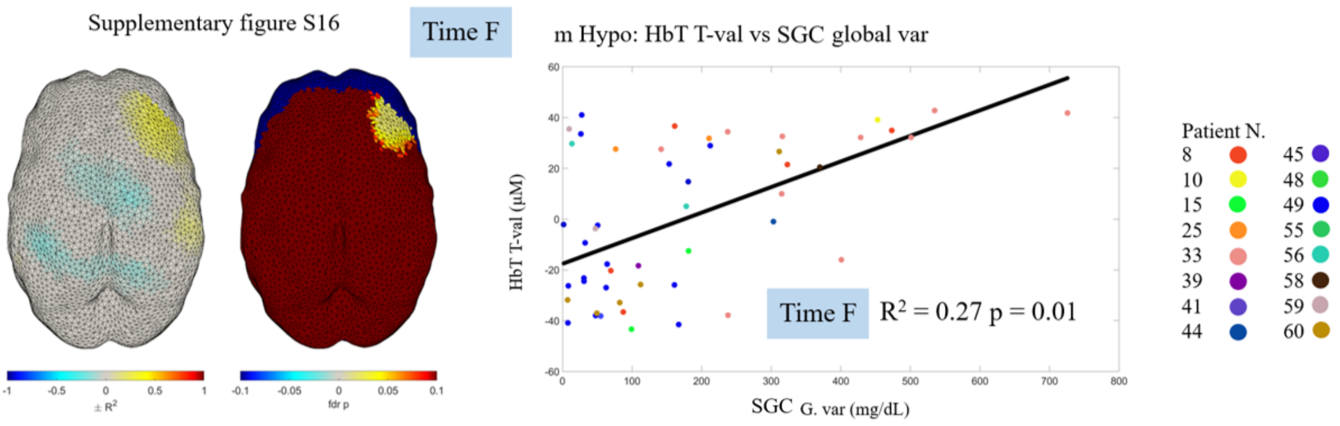


S16: On the left of the figure, the $R^2$ on the left and the FDR-corrected p-values on the right, for each node on the cortex displaying the correlation between HbT T-val and SGC global var for m-hypo events (52 events) in time window F. At the bottom of the figure is the correlation between the two metrics across all events for the cortical node with the highest $R^2$ value.

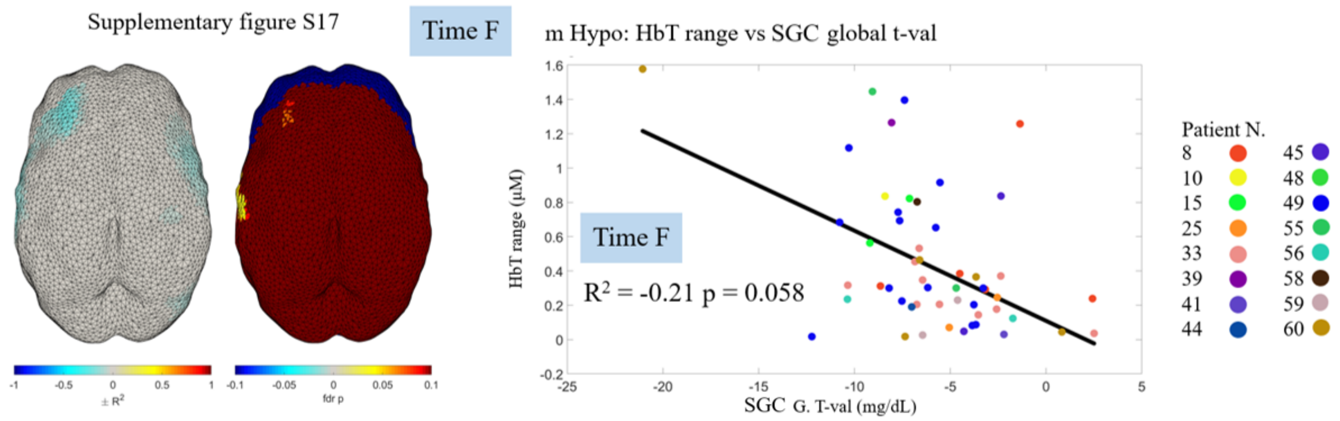


S17: On the left of the figure, the $R^2$ on the left and the FDR-corrected p-values on the right, for each node on the cortex displaying the correlation between HbT range and SGC global t-val for m-hypo events (52 events) in time window F. At the bottom of the figure is the correlation between the two metrics across all events for the cortical node with the highest $R^2$ value.

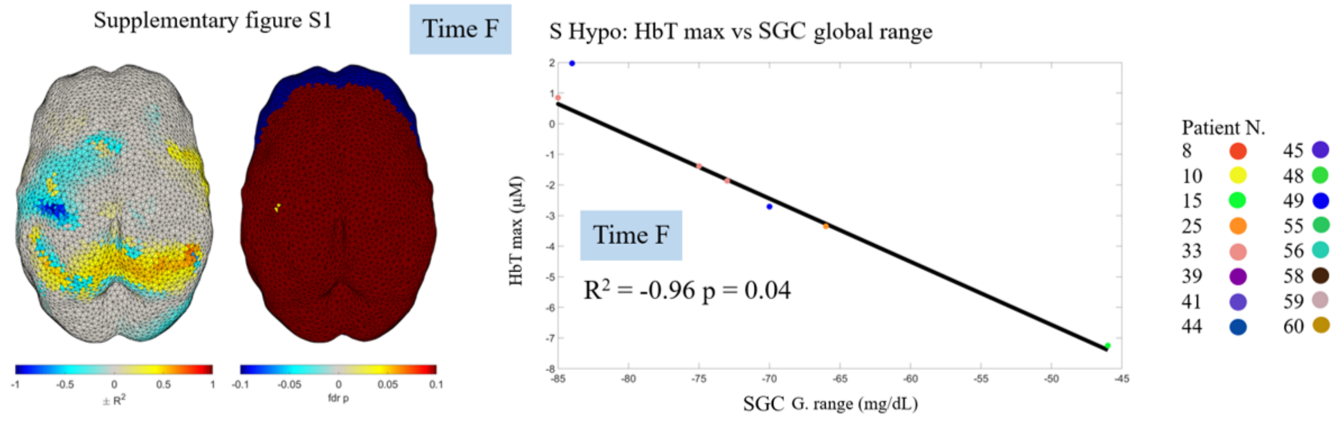


S18: On the left of the figure, the $R^2$ on the left and the FDR-corrected p-values on the right, for each node on the cortex displaying the correlation between HbT max and SGC global range for S-hypo events (7 events) in time window F. At the bottom of the figure is the correlation between the two metrics across all events for the cortical node with the highest $R^2$ value.
